# ClinGen Hemoglobinopathy VCEP ACMG/AMP specifications for *HBB* and *HBA2* variant interpretation in thalassemia

**DOI:** 10.64898/2026.10.30.26366794

**Authors:** Coralea Stephanou, Irene Fylaktou, Hashim Halim-Fikri, Tamara T. Koopmann, Landry E Nfonsam, Celeste Bento, Cornelis L Harteveld, Marina Kleanthous, Joanne Traeger-Synodinos, Kyriaki Michailidou, Thessalia Papasavva, Bin Alwi Zilfalil, Justyne E Ross, John S Waye, Carsten W Lederer, Petros Kountouris, the ClinGen Hemoglobinopathy Variant Curation Expert Panel

## Abstract

**Background:** The ClinGen Hemoglobinopathy Variant Curation Expert Panel (VCEP) was established in 2019 to develop disease-specific adaptations of the American College of Medical Genetics and Genomics/Association for Molecular Pathology (ACMG/AMP) sequence variant interpretation framework for hemoglobinopathies. This study describes the development, validation, and ClinGen approval of the first disease-specific ACMG/AMP specifications for autosomal recessive thalassemia related to *HBB* and *HBA2,* providing a standardized framework for globin gene variant interpretation.

**Methods:** Disease-specific ACMG/AMP rule specifications were developed in accordance with ClinGen procedures and Variant Classification Working Group recommendations and evaluated through pilot curation of representative *HBB* and *HBA2* variants using the ClinGen Variant Curation Interface (VCI), with independent biocurator assessment, expert review, and consensus approval.

**Results:** A total of 34 disease-specific evidence specifications were developed across the two gene-disease pairs, encompassing 22 ACMG/AMP evidence criteria and one modification to the ACMG/AMP evidence-combining rules. Pilot application to 33 representative variants (20 *HBB*, 13 *HBA2*) demonstrated high concordance with established pathogenic and benign classifications while providing a standardized framework for resolving variants with conflicting or limited evidence. Following pilot validation, the specifications received ClinGen approval, and all curated variants with supporting evidence were disseminated through the ClinGen Evidence Repository (ERepo) and ClinVar.

**Conclusions:** The ClinGen Hemoglobinopathy VCEP specifications provide the first ClinGen-approved disease-specific ACMG/AMP rule specifications for *HBB* and *HBA2* variants related to thalassemia. These specifications establish a robust and transparent framework for harmonized globin variant interpretation and provide a foundation for sustained expert curation, continued refinement through emerging evidence, and expansion to additional hemoglobinopathy gene-disease pairs.

## Background

Hemoglobinopathies are among the most prevalent hereditary diseases in the world. They comprise a group of inherited blood disorders affecting hemoglobin, the oxygen-carrying protein of red blood cells, and are broadly categorized into two main groups: structural hemoglobin variants, which alter the structure or functional properties of globin proteins, and thalassemia syndromes, characterized by reduced or absent globin chain synthesis [1,2].

Among hemoglobinopathies, thalassemia syndromes constitute a substantial global health burden. A recent analysis of Global Burden of Disease (GBD) 2021 data estimated that 1.31 million individuals are living with thalassemia [3]. The underlying carrier burden is considerably greater, with an estimated 5–20% of the global population carrying one or more α-thalassemia variants [4] and approximately 1.5% carrying a β-thalassemia-associated variant [5]. Although α-thalassemia is considerably more common, β-thalassemia accounts for a disproportionate burden of severe disease, with more than 40,000 affected births annually, nearly two-thirds of which represent transfusion-dependent thalassemia [3,6]. The global distribution of thalassemia is characterized by marked regional differences in allele frequencies, with substantial differences observed even across relatively small geographic regions. These patterns have been largely shaped by historical malaria-driven balancing selection, which favored heterozygous carriers and contributed to the high prevalence of specific pathogenic globin variants in affected populations [1].

The α– and β-globin genes are organized into two highly conserved, developmentally regulated gene clusters located on chromosome 16 (α-globin locus; Accession: NG_000006) and 11 (β-globin locus; Accession: NG_000007), respectively. The α-globin locus contains the highly sequence-similar *HBA1* and *HBA2*, while the β-globin locus comprises *HBB*, alongside embryonic and fetal globin genes, all tightly coordinated by upstream regulatory locus control regions [7]. Pathogenic variants affecting these loci variously disrupt globin chain synthesis, protein stability, oxygen affinity, tetramer assembly, and erythrocyte survival, giving rise to the diverse clinical spectrum of hemoglobinopathies [8].

The molecular basis of thalassemia is highly heterogeneous and includes both single nucleotide variants (SNVs) and copy number variants (CNVs) affecting globin genes and their regulatory elements, with more than 2000 mutant alleles reported to date [9,10]. Individuals normally carry four α-globin genes, one copy each of *HBA1* and *HBA2* on both chromosomes 16 (αα/αα), but only two β-globin genes, one *HBB* copy on each chromosome 11 (β/β). This difference contributes to the distinct genotype-phenotype relationships of α− and β-thalassemia. In α-thalassemia, loss of one α-globin gene results in a clinically silent carrier state (−α/αα), whereas loss of two genes causes α-thalassemia trait (−−/αα or −α/−α).

Inactivation of three α−globin genes leads to moderate-to-severe HbH disease (−−/−α), while absence of all four genes (−−/−−) results in α-thalassemia major (hydrops fetalis), a condition that is typically lethal in utero or shortly after birth [4]. Conversely, β-thalassemia results from pathogenic variants affecting one or both *HBB* alleles that reduce or abolish β-globin synthesis primarily through defects in transcription, RNA processing, translation, or globin chain stability. Depending on the extent of residual β-globin production, alleles are traditionally classified as mild (β^++^), moderate (β^+^), or complete loss of function (β^0^). Clinical severity is determined by the combination of the two inherited alleles, ranging from clinically asymptomatic carriers to severe transfusion-dependent disease with multisystem complications [11]. The extensive allelic heterogeneity, the involvement of two globin gene loci in disease manifestation, and the influence of modifier genes contribute substantially to the complexity of molecular diagnosis and clinical variant interpretation [12].

To standardize variant interpretation and minimize laboratory discordance, the American College of Medical Genetics and Genomics and the Association for Molecular Pathology (ACMG/AMP) published consensus recommendations in 2015 for sequence variants in Mendelian disorders [13]. This framework utilizes a five-tier classification system comprising benign (B), likely benign (LB), variants of uncertain significance (VUS), likely pathogenic (LP) and pathogenic (P), derived from a semiquantitative integration of population, computational, functional, segregation, and allelic data. Individual evidence codes are assigned specific strengths, ranging from supporting to very strong (pathogenic) or stand-alone (benign). While these guidelines provide a robust foundation, their broad design necessitates gene– and disease-specific adaptations to account for disease-specific molecular and clinical characteristics. This is particularly relevant for the globin gene loci, where the application of population-based, functional, and case-level evidence must account for unique molecular mechanisms, genotype-phenotype relationships, and population-specific allele distributions.

Accordingly, the Clinical Genome Resource (ClinGen) [14] established the Variant Curation Expert Panel (VCEP) framework to support the development of gene– and disease-specific ACMG/AMP specifications via expert consensus and systematic evaluation. The ClinGen Hemoglobinopathy VCEP was formed as an international collaborative effort through the ITHANET portal [9,10] and the Global Globin Network of the Human Variome Project [15] to develop standardized specifications for the interpretation of globin gene variants associated with hemoglobinopathies [16]. The present study reports the finalized ClinGen-approved ACMG/AMP specifications for the interpretation of SNVs in *HBB* and *HBA2* related thalassemia. These specifications represent the first ClinGen-approved ACMG/AMP recommendations developed by the Hemoglobinopathy VCEP and correspond to the initial *HBB* and *HBA2* gene-disease pairs formally evaluated under the ClinGen Gene Curation framework [17]. Developed through international expert consensus and informed by extensive pilot variant curation, these specifications provide a standardized framework for the U.S. Food and Drug Administration (FDA)-recognized globin variant interpretation aimed at improving consistency across laboratories, resolving conflicting classifications, and reducing the burden of variants of uncertain significance in hemoglobinopathy diagnostics.

## Methods

Figure 1 provides an overview of the development, pilot validation, and ClinGen approval pathway for the Hemoglobinopathy VCEP specifications. The methodology underpinning each step is described below.

**Figure 1.**
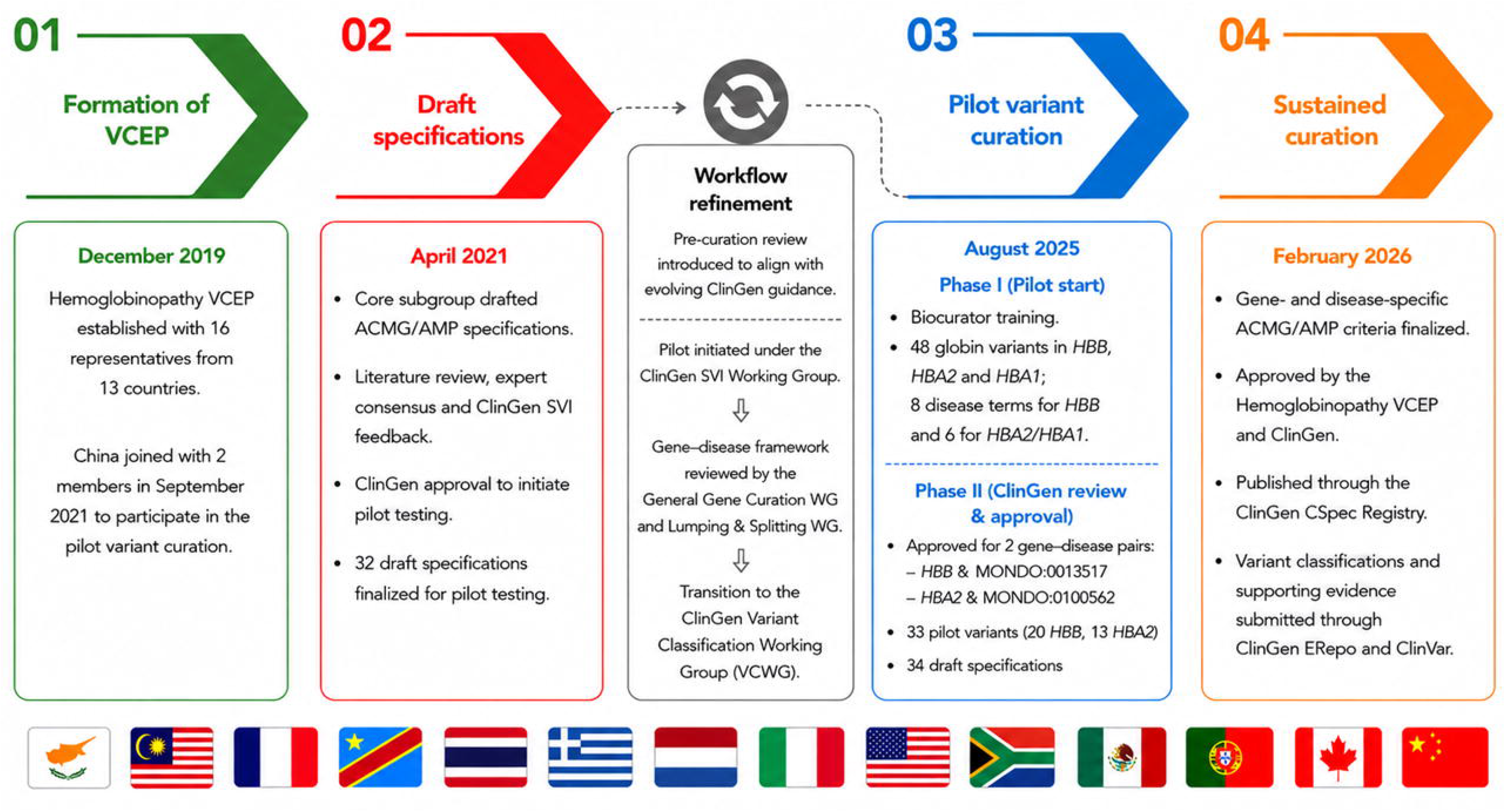
Development, validation, and ClinGen approval of the Hemoglobinopathy VCEP specifications. The figure summarizes the sequential workflow leading to the development, pilot testing, refinement, and ClinGen approval of the Hemoglobinopathy VCEP gene– and disease-specific ACMG/AMP variant classification specifications. VCEP, Variant Curation Expert Panel; SVI, Sequence Variant Interpretation; VCWG, Variant Curation Working Group

### Panel composition and scope

The Hemoglobinopathy VCEP (ClinGen affiliation ID 50052, https://www.clinicalgenome.org/affiliation/50052/) was established in December 2019 to develop gene– and disease-specific adaptations of the ACMG/AMP sequence variant interpretation framework [13] for hemoglobinopathies (Step 1). At its inception, the VCEP comprised 26 experts in hemoglobinopathies from 13 countries across Europe, Asia, Africa, and North/Central America, with expertise spanning clinical and laboratory genetics, molecular diagnostics, hematology, bioinformatics, database curation, population genetics, and translational research. A center in China joined with two members during the pilot testing of the draft specifications. All members disclosed potential conflicts of interest in accordance with ClinGen requirements.

### Rule specification process

A core subgroup representing diverse expertise within the panel convened regularly to review the applicability, strength modifications, and disease-specific considerations of the ACMG/AMP criteria for SNVs in *HBB* (HGNC: 4827), *HBA2* (HGNC: 4824), and *HBA1* (HGNC: 4823) associated with hemoglobinopathies. Initial specifications were developed based on published evidence and expert review, followed by iterative refinement and consensus approval by the full VCEP through teleconferences and e-mail correspondence. Published recommendations and feedback from the ClinGen Sequence Variant Interpretation (SVI) Working Group, now part of the ClinGen Variant Classification Working Group (VCWG), were incorporated through multiple rounds of review and revision. Criteria deemed not applicable were excluded, whereas others underwent gene– or disease-specific adaptation. Computational evidence specifications were informed by an independent benchmarking study of in silico prediction tools across globin gene variant types [18]. These efforts culminated in ClinGen approval of the ACMG/AMP specifications for pilot testing in April 2021 (Step 2). The framework comprised 34 specified evidence codes, including criteria applied at different strengths according to the quantity and quality of available evidence. Criteria were combined according to the original ACMG/AMP framework to derive variant classifications [13]. The complete ClinGen-approved specifications are publicly available through the ClinGen Criteria Specification Registry (CSpec, https://cspec.genome.network), which is updated periodically. Readers should refer to the online registry for the most current versions of the specifications, including *HBB* and beta-thalassemia HBB/LCRB (GN170; MONDO:0013517), and *HBA2* and HBA2-related alpha thalassemia spectrum (GN173; MONDO:0100562).

### Pilot variant selection

A final pilot set of 20 *HBB* and 13 *HBA2* variants with thalassemia-associated annotations in ClinVar [19] and ITHANET [9] was selected to validate the specified ACMG/AMP criteria. Variants represented diverse molecular consequences, including promoter and 5′UTR variants affecting transcription; consensus and cryptic splice-site, polyadenylation signal, and 3′UTR variants affecting mRNA processing; initiation codon, nonsense, and antitermination variants affecting translation; missense variants; and other intronic variants. Loss-of-function variants were intentionally limited because PVS1 alone may provide Very Strong pathogenic evidence, while variants with uncertain classifications were included to assess criterion applicability in the presence of insufficient and/or conflicting evidence. The pilot set was thus designed to maximize assessment of ACMG/AMP criteria across diverse variant contexts and classification outcomes.

### Pilot variant curation and review

Pilot variants were independently reviewed by pairs of trained biocurators responsible for literature review, evidence collection, and data aggregation, under the coordination of a biocurator trainer. Curated evidence and preliminary assessments were compared and discussed in dedicated biocurator meetings, after which an assigned biocurator formalized each curation in the ClinGen Variant Curation Interface (VCI) [20], including evidence documentation, application of the specified ACMG/AMP criteria, standardized evidence summaries, and provisional classification. Publicly available evidence was supplemented, where available, by anonymized case-level observations contributed by panel member laboratories. VCI-documented evidence and provisional classifications were made available to the full panel for expert review prior to dedicated review meetings. Panel-reviewed classifications required agreement from at least three core approval members, with discrepancies resolved through expert consensus discussions. The pilot phase involved six trained biocurators and 13 panel experts.

### Final *HBB* and *HBA2* specifications

During pilot testing and review, evolving ClinGen VCEP procedures incorporated gene-disease validity assessments [21], standardized disease entity definitions [22], and lumping and splitting recommendations [23]. The specifications were aligned with these requirements without altering their underlying evidence logic. Although pilot testing initially encompassed multiple hemoglobinopathy gene-disease pairs, final validation focused on *HBB* for beta-thalassemia HBB/LCRB (MONDO:0013517) and *HBA2* for HBA2-related alpha thalassemia spectrum (MONDO:0100562), both under an autosomal recessive inheritance model (HP:0000007). A disease term selection framework and variant-type-based decision flowchart were developed to support consistent variant curation (Supplementary Figure 1). Final SVI recommendations and subsequent ClinGen VCEP-wide guidance, including the transition to gnomAD v4 for population data, were incorporated before the specifications were resubmitted for approval. The finalized gene– and disease-specific specifications for *HBB* and *HBA2* completed ClinGen review in August 2025 (Step 3).

### Public deposition of variant classifications

Following completion of the ClinGen approval process in February 2026, final variant classifications and supporting evidence were submitted to the ClinGen Evidence Repository (ERepo, https://erepo.genome.network/evrepo/ui) and subsequently deposited in ClinVar under the Hemoglobinopathy VCEP affiliation as FDA-recognized expert panel submissions (https://www.ncbi.nlm.nih.gov/clinvar/submitters/510550/) (Step 4).

## Results

### Summary of criterion specifications

The Hemoglobinopathy VCEP developed specifications for 22 ACMG/AMP criteria, while six criteria were excluded from both gene-disease frameworks, either due to limited applicability in globin gene variant interpretation (PP2, PP4, BP1, BP3) or following recommendations from the ClinGen SVI Working Group (PP5, BP6) [24]. The resulting framework comprises 34 individual evidence specifications. For *HBB*–β-thalassemia, 14 criteria received gene– and/or disease-specific modifications (PVS1, PS2, PS3, PS4, PM1, PM2, PM5, PP1, PP3, BA1, BS1, BS2, BS3, BP4), 8 were adapted through strength-level modifications (PVS1, PS3, PS4, PM2, PM3, PP1, BS2, BS3), and 7 followed the original ACMG/AMP recommendations unchanged (PS1, PM4, PM6, BS4, BP2, BP5, BP7). A nearly identical framework was developed for *HBA2*–α-thalassemia, differing only in two aspects: BP5 was considered not applicable, and an additional specification, PS1_Moderate, was introduced to account for amino acid substitutions previously established as pathogenic in the paralogous *HBA1*. A complete summary of the criteria specifications and their respective designations across both gene-disease pairs is provided in Table 1. The modified ACMG/AMP evidence-combining rules adopted by the Hemoglobinopathy VCEP are summarized in Table 2, including recognition of ≥2 independent Very Strong criteria (PVS1, PM3 and PS4 at Very Strong strength) as sufficient for a pathogenic classification.

**Table 1.** Hemoglobinopathy VCEP specifications of the ACMG/AMP criteria for HBB and HBA2 variant classification. Original ACMG/AMP criteria and their corresponding Hemoglobinopathy VCEP specifications are presented together with the applicable gene–disease pair(s). Modification type indicates whether criteria underwent gene– and/or disease-specific adaptation, modification of evidence strength, remained unchanged from the original ACMG/AMP recommendations, or were removed as not applicable. Full specifications available from the CSpec: HBB, https://cspec.genome.network/cspec/ed/svi/doc/GN170; HBA2, https://cspec.genome.network/cspec/ed/svi/doc/GN173

| ACMG/AMP criteria | Modification Type | Applies to | Criteria Description | Criteria Specification |
|---|---|---|---|---|
| **PVS1** | Disease-specific, Gene-specific | HBB/MONDO:0013517; HBA2/MONDO:0100562 | Null variant (nonsense, frameshift, canonical +/-1 or 2 splice sites, initiation codon, single or multi-exon deletion) in a gene where loss of function (LOF) is a known mechanism of disease.<br>Caveats include i) LOF variants at the extreme 3’ end of a gene and ii) splice variants predicted to lead to exon skipping but leave the remainder of the protein intact. | Loss-of-function has been established as a disease mechanism for hemoglobinopathies. |
|  | Strength | HBB/MONDO:0013517; HBA2/MONDO:0100562 | **PVS1_Strong** and **PVS1_Moderate** are assigned according to predicted variant consequence, predicted NMD status, and variant location. | Hemoglobinopathy VCEP PVS1 decision tree (Figure 2) |
| **PS1** | No change | HBB/MONDO:0013517; HBA2/MONDO:0100562 | Same amino acid change as a previously established pathogenic variant regardless of nucleotide change. | A variant classification must be established by a VCEP classification (CSpec) or VCEP consensus recommendation. |
|  | Strength | HBA2/MONDO:0100562 | **PS1_Moderate:** The same change of an amino acid shown to be pathogenic in a paralogue gene. |  |
| **PS2** | Disease-specific, Gene-specific | HBB/MONDO:0013517; HBA2/MONDO:0100562 | De novo (both maternity and paternity confirmed) in a patient with the disease or in a phenotypic trait individual and no family history.<br>Note: Confirmation of paternity only is insufficient. Egg donation, surrogate motherhood, errors in embryo transfer, etc. can contribute to non-maternity. | Both maternal and paternal sample must be tested and shown to be the biological parents of the affected individual. Otherwise, identity is assumed, and not confirmed, and PM6 applies.<br>Only applicable in the absence of any other established pathogenic variants. If other suspicious variants are present, then PM6 should be used instead. Should not be used in combination with PM6.<br>Definition of trait phenotype is provided in Table 3. |
| **PS3_Supporting** | Disease-specific, Gene-specific, Strength | HBB/MONDO:0013517; HBA2/MONDO:0100562 | In vitro or in vivo functional studies supportive of a damaging effect on the gene, gene product, expression levels and protein function. | Approved functional studies include: Hemoglobin stability test; Biosynthesis assay; Hemoglobin electrophoresis, HPLC; in vitro splicing assay; in vitro cell-based assay. |
| **PS4** | Disease-specific, Gene-specific, | HBB/MONDO:0013517; HBA2/MONDO:0100562 | The prevalence of the variant in affected individuals is significantly increased compared with the prevalence in controls. |  |
|  | Strength | HBB/MONDO:0013517; HBA2/MONDO:0100562 | **PS4_Very Strong** (≥16 points), **PS4** (3.5–15.99 points), **PS4_Moderate** (1.5–3.49 points), **PS4_Supporting** (0.5–1.49 points) | Strength evidence is assigned using a point-based framework (Table 3). |
| **PM1** | Disease-specific, Gene-specific | HBB/MONDO:0013517; HBA2/MONDO:0100562 | Located in a mutational hot spot and/or critical and well-established functional domain (e.g. active site of an enzyme) without benign variation. | **HBB/MONDO:0013517:** HBB promoter TATAA box (-30 to -26 from transcription initiation; c.-80 to c.-76); Poly(A) signal AATAAA sequence<br>**HBA2/MONDO:0100562:** AHSP binding (α32R, α104H, α118F, α120P); Poly(A) signal AATAAA sequence |
| **PM2_Supporting** | Disease-specific, Gene-specific, Strength | HBB/MONDO:0013517; HBA2/MONDO:0100562 | Allele frequency <0.0001 (0.01%) in gnomAD, with sufficient sequence coverage (≥20x) at the variant position. | Position of reported variant must have sufficient coverage (>= 20x) in the population database. |
| **PM3** | Disease-specific, Gene-specific | HBB/MONDO:0013517; HBA2/MONDO:0100562 | For recessive disorders, detected in trans with a pathogenic or likely pathogenic variant in an affected patient. | Both variants must meet PM2_Supporting specification, i.e. be sufficiently rare, unless they are in the exception list (Supplementary Table 1), or present in gnomAD with <20x genomic coverage.<br>Phase is confirmed in trans by testing both or one parent, and pathogenicity of the variant on the other allele must be established by a VCEP classification (CSpec) or VCEP consensus recommendation. |
|  | Strength | HBB/MONDO:0013517; HBA2/MONDO:0100562 | **PM3_Very Strong**, **PM3_Strong**, **PM3_Supporting** | Strength assignment follows ClinGen SVI PM3 recommendations (https://clinicalgenome.org/site/assets/files/3717/svi_proposal_for_pm3_criterion_-_version_1.pdf) |
| **PM4** | No change | HBB/MONDO:0013517; HBA2/MONDO:0100562 | Protein length changes as a result of in-frame deletions/insertions in a non-repeat region or stop-loss variants. | Should not be used, if PVS1 has been applied.<br>**HBA2/MONDO:0100562**: Relevant for stop codon mutations leading to elongated α-chains (e.g., Hb Constant Spring) and in-frame deletions of a complete codon (leading to deletion of a single amino acid in an otherwise functional hemoglobin variant). |
| **PM5** | Disease-specific, Gene-specific | HBB/MONDO:0013517; HBA2/MONDO:0100562 | Novel missense change at an amino acid residue where a different missense change determined to be pathogenic has been seen before, or base change at a non-coding position and previous pathogenic mutation has been seen before.<br>Examples: Arg156His is pathogenic; now you observe Arg156Cys.<br>Non-coding variant g.15300G>C is pathogenic, now you observe g. 15300G>A. | Beware of changes that impact splicing rather than at the amino acid/protein level. Assess potential for creation of exonic splicing event using in silico splicing prediction tools, as described in PP3/BP4.<br>A previously established pathogenic variant must a VCEP classification (CSpec) or VCEP consensus recommendation. |
| **PM6** | No change | HBB/MONDO:0013517; HBA2/MONDO:0100562 | Assumed de novo without confirmed maternity and paternity testing. | In contrast to PS2, identity testing was NOT performed in parental samples to confirm identity, which is assumed. Should not be used in combination with PS2. |
| **PP1** | Disease-specific, Gene-specific, | HBB/MONDO:0013517; HBA2/MONDO:0100562 | Co-segregation with disease in multiple affected family members in a gene definitively known to cause the disease. | Caution needed when counting segregations in the presence of other possible disease-causing variants. Compound heterozygous individuals are counted only if phase is confirmed to be in trans. |
|  | Strength | HBB/MONDO:0013517; HBA2/MONDO:0100562 | **PP1_Strong** (>2.1 LOD score), **PP1_Moderate** (>1.5 LOD score), **PP1** (>0.9 LOD score) | LOD score for recessive disorder can be estimated from the tables in Supplementary, based on the number of affected and unaffected individuals. |
| **PP2** | Removed |  | Missense variant in a gene that has a low rate of benign missense variation and where missense variants are a common mechanism of disease. | Globin genes are affected by both pathogenic and benign missense variants. |
| **PP3** | Disease-specific, Gene-specific | HBB/MONDO:0013517; HBA2/MONDO:0100562 | Multiple lines of computational evidence support a deleterious effect on the gene or gene product (conservation, evolutionary, splicing impact, etc.).<br>For missense: REVEL score > 0.8 OR SpliceAI > 0.3. If REVEL score is not available, use CADD PHRED score > 23.5<br>For non-coding, synonymous, in-frame indels, stop-lost: CADD PHRED score > 12 OR SpliceAI DS > 0.3 | Should not be used for LOF variants considered in PVS1. |
| **PP4** | Removed |  | Patient’s phenotype or family history is highly specific for a disease with a single genetic etiology. | Phenotype-based evidence is already incorporated into PS4 (observation in heterozygous) and PM3 (allelic observations in trans) |
| **PP5** | Removed |  | Reputable source recently reports variant as pathogenic, but the evidence is not available to the laboratory to perform an independent evaluation. | SVI recommendations |
| **BA1** | Disease-specific, Gene-specific | HBB/MONDO:0013517; HBA2/MONDO:0100562 | Allele frequency ≥0.005 (0.5%) in a studied general population with ≥2000 alleles and variant present in ≥5 alleles. | Use the Filtering AF, such as Popmax FAF from gnomAD. If the variant is present at high frequency in the non-continental population (e.g., Ashkenazi Jews), you can calculate the filtering allele frequency using a 95% confidence interval by selecting "Inverse AF" at http://cardiodb.org/allelefrequencyapp/<br>Assumptions to calculate this threshold for:<br>**HBB/MONDO:0013517**<br>●Prevalence: 1/1000 (the highest reported prevalence of the disease)<br>●Genetic heterogeneity: 100% (only one gene involved in disease causality)<br>●Allelic heterogeneity: 15% (variants in the exclusion list are not considered)<br>●Penetrance: 90% (to account for silent variants)<br>**HBA2/MONDO:0100562**<br>●Prevalence: 1/1000 (the highest reported prevalence of the disease)<br>●Genetic heterogeneity: 75% (HBA2 is affected in around 75% of pathogenic variants, from IthaGenes database)<br>●Allelic heterogeneity: 15% (i.e. a single variant responsible for all of cases)<br>●Penetrance: 80% (to account for silent variants and for underdiagnosis of Hb H disease)<br>Variants in Supplementary Table 1 are excluded from this criterion. |
| **BS1** | Disease-specific, Gene-specific | HBB/MONDO:0013517; HBA2/MONDO:0100562 | Allele frequency ≥0.001 (0.1%) in a studied general population with ≥2000 alleles and variant present in ≥5 alleles. | Use the Filtering AF, such as Popmax FAF from gnomAD. If the variant is present at high frequency in the non-continental population (e.g. Ashkenazi Jews), you can calculate the filtering allele frequency using a 95% confidence interval by selecting "Inverse AF" at http://cardiodb.org/allelefrequencyapp/<br>Assumptions to calculate this threshold:<br>**HBB/MONDO:0013517**<br>●Prevalence: 1/3000 (the highest reported prevalence of the disease)<br>●Genetic heterogeneity: 100% (only one gene involved in disease causality)<br>●Allelic heterogeneity: 5% (variants in the exclusion list are not considered)<br>●Penetrance: 90% (to account for silent variants)<br>**HBA2/MONDO:0100562**<br>●Prevalence: 1/3000 (the highest reported prevalence of the disease)<br>●Genetic heterogeneity: 75% (HBA2 is affected in around 75% of pathogenic variants – from IthaGenes database)<br>●Allelic heterogeneity: 5% (variants in the exclusion list are not considered)<br>●Penetrance: 80% (to account for silent variants and for underdiagnosis of Hb H disease)<br>Variants in Supplementary Table 1 are excluded from this criterion. |
| **BS2** | Disease-specific, Gene-specific, | HBB/MONDO:0013517; HBA2/MONDO:0100562 | Two independent occurrences in individuals (asymptomatic or with trait phenotype) for a recessive (homozygous or compound heterozygous) disorder, with full penetrance expected at early age. | Normal hematological values are provided in Supplementary<br>The healthy individual must be well-phenotyped/well-documented to rule out any mild symptoms and ensure that the individual is unaffected. A full-blood count and Hb characterization are required to exclude variants and/or abnormal quantities.<br>In compound heterozygous individuals, only established pathogenic variants should be considered (i.e. supported by a VCEP classification (CSpec) or VCEP consensus recommendation).<br>Applies to subjects over 2 years of age.<br>**BS2**<br>**HBB/MONDO:0013517:** Only applicable if no coinheritance is detected of an HBA1/HBA2 pathogenic variant.<br>**HBA2/MONDO:0100562:** Only applicable if no coinheritance is detected of an HBB pathogenic variant.<br>**BS2_P**: Only applicable if no well-established disease-modifying mutations are detected. |
|  | Strength | HBB/MONDO:0013517; HBA2/MONDO:0100562 | **BS2_Supporting:** Observation in one individual (asymptomatic or with trait phenotype) for a recessive (homozygous or compound heterozygous) disorder, with full penetrance expected at early age. |  |
| **BS3** | Disease-specific, Gene-specific, Strength | HBB/MONDO:0013517; HBA2/MONDO:0100562 | In vivo or in vitro functional studies show no damaging effect on gene, gene product, expression levels and protein function. | Functional studies demonstrate no damaging effect using approved assays (hemoglobin stability testing, biosynthesis assay, electrophoresis/HPLC, in vitro splicing assay, or cell-based assays). Thresholds are provided in Supplementary Table 2.. |
| **BS4** | No change | HBB/MONDO:0013517; HBA2/MONDO:0100562 | Lack of segregation in affected members of a family. |  |
| **BP1** | Removed |  | Missense variant in a gene for which primarily truncating variants are known to cause disease. | Globin genes are affected by both pathogenic and benign missense variants, in addition to truncating variants. |
| **BP2** | No change |  | Observed in cis with a pathogenic variant in any inheritance pattern. | Do not use when the variant has only ever been observed in cis with a pathogenic variant as its significance/severity in isolation is unknown.<br>Only applies when the phenotype is not more severe than when either of the two variants are seen in isolation.<br>Use only if in cis with variants classified as pathogenic are supported by a VCEP classification (CSpec) or VCEP consensus recommendation. |
| **BP3** | Removed |  | In frame-deletions/insertions in a repetitive region without a known function. | Globin genes do not contain a repetitive region without known function |
| **BP4** | Disease-specific, Gene-specific | HBB/MONDO:0013517; HBA2/MONDO:0100562 | Multiple lines of computational evidence suggest no impact on gene or gene product (conservation, evolutionary, splicing impact, etc.) | For missense: REVEL score < 0.7 AND SpliceAI DS ≤ 0.3 (if REVEL score is not available, use CADD PHRED score ≤ 20, instead)<br>For non-coding, synonymous, in-frame indels, stop-lost: CADD PHRED score ≤ 11 AND SpliceAI DS ≤ 0.3 |
| **BP5** | Disease-specific, Gene-specific | HBB/MONDO:0013517 | Variant found in a case with an alternate molecular basis for disease. | This is about phenotypes that are COMPLETELY explained by variants in different genes. Specifically:<br>●duplication of the α-locus can result in β-thalassemia intermedia phenotype<br>●variants in SUPT5H that can cause β-thalassemia phenotype |
| **BP6** | Removed |  | Reputable source recently reports variant as benign, but the evidence is not available to the laboratory to perform an independent evaluation. | SVI recommendations |
| **BP7** | No change |  | A synonymous (silent) variant for which splicing prediction algorithms predict no impact to the splice consensus sequence nor the creation of a new splice site AND the nucleotide is not highly conserved. | Synonymous variant with no impact on splicing (SpliceAI ≤0.3) AND GERP++ <0 for conservation. |

**Table 2.** Hemoglobinopathy VCEP evidence combinations for variant classification. The table summarizes the combinations of pathogenic and benign evidence used to assign pathogenic (P), likely pathogenic (LP), benign (B), likely benign (LB), and variant of uncertain significance (VUS) classifications. The only modification to the original ACMG/AMP framework is the recognition of ≥2 independent Very Strong pathogenic criteria as sufficient to support a pathogenic classification.

| Classification | Evidence combinations |
|---|---|
| **Pathogenic (P)** | ≥2 Very Strong **(Hemoglobinopathy VCEP modification)** |
|  | 1 Very Strong + ≥1 Strong |
|  | 1 Very Strong + ≥2 Moderate |
|  | 1 Very Strong + 1 Moderate + 1 Supporting |
|  | 1 Very Strong + ≥2 Supporting |
|  | ≥2 Strong |
|  | 1 Strong + ≥3 Moderate |
|  | 1 Strong + 2 Moderate + ≥2 Supporting |
|  | 1 Strong + 1 Moderate + ≥4 Supporting |
| **Likely Pathogenic (LP)** | 1 Very Strong + 1 Moderate |
|  | 1 Strong + 1-2 Moderate |
|  | 1 Strong + ≥2 Supporting |
|  | ≥3 Moderate |
|  | 2 Moderate + ≥2 Supporting |
|  | 1 Moderate + ≥4 Supporting |
| **Likely Benign (LB)** | 1 Strong + 1 Supporting |
|  | ≥2 Supporting |
| **Benign (B)** | 1 Stand-alone |
|  | ≥2 Strong |
| **Variant of Uncertain Significance (VUS)** | Criteria for Pathogenic/Likely Pathogenic or Benign/Likely Benign are not met, **or** pathogenic and benign evidence are contradictory. |

### Population data (BA1, BS1, PM2_Supporting)

#### BA1, BS1: population frequency thresholds

BA1 (benign stand-alone) and BS1 indicate that a variant is too frequent in the general population to be causative of disease. Disease-specific thresholds were derived using the Whiffin/Ware allele frequency framework and calculator [25], incorporating conservative assumptions regarding disease prevalence, penetrance, and genetic and allelic heterogeneity. For BA1, calculations assumed the highest reported disease prevalence (1 in 1,000 individuals) [26], penetrance values of 90% for *HBB* and 80% for *HBA2* (to account for silent variants and, for *HBA2*, potential underdiagnosis of HbH disease), genetic heterogeneity of 100% for *HBB* and 75% for *HBA2* (reflecting the estimated contribution of *HBA2* to pathogenic α-thalassemia variants curated in the IthaGenes globin variant database [9]), and a maximum allelic contribution of 15% (i.e., assuming no single pathogenic variant accounts for more than 15% of disease alleles). These assumptions resulted in a BA1 threshold of ≥0.005 (0.5%). For BS1, a more stringent disease prevalence of 1 in 3,000 individuals and a maximum allelic contribution of 5% were assumed, yielding a threshold of ≥0.001 (0.1%). Consistent with SVI recommendations, both criteria additionally required the variant to be observed in at least five alleles within a reference dataset comprising a minimum of 2,000 alleles [27]. Population frequencies were assessed using the GroupMax Filtering Allele Frequency (FAF; 95% confidence interval) from gnomAD v4, which represents the highest filtering allele frequency among non-bottlenecked ancestry groups.

To avoid inappropriate application of population frequency criteria because of malaria-driven selection or founder effects, the VCEP established a population-frequency exclusion list using allele frequency data curated in IthaMaps, which compiles country– and region-specific frequencies for globin gene variants worldwide [9,10]. Variants included in the exclusion list were exempt from BA1 and BS1 application and from the PM2_Supporting rarity requirement when evaluating PM3 (Supplementary Table 1).

#### PM2_Supporting: rarity in population databases

Consistent with SVI recommendations [28], the VCEP adopted PM2 at the Supporting strength level (PM2_Supporting) for variants that are absent or rare in population reference datasets. The allele frequency threshold was set at <0.0001 (0.01%), one order of magnitude below the BS1 threshold of ≥0.001 (0.1%) [29]. In accordance with the approved specifications, PM2_Supporting is applied using gnomAD v4 when a variant has a total allele frequency <0.0001 (0.01%), read depth ≥20, and passes gnomAD quality control filters.

### Variant type and location (PVS1, PS1, PM1, PM4, PM5)

#### PVS1: null variants

Loss of function (LoF) is an established disease mechanism for thalassemia, supported by the absence of benign or likely benign null variants in *HBB* and *HBA2* in ClinVar and by likelihood ratio analyses demonstrating a consistent association between null variants and pathogenicity. Consequently, PVS1 is applicable to sequence variants (SNVs and short indels) predicted to abolish normal gene function, including nonsense, frameshift, canonical ±1 and ±2 splice sites, and initiation codon variants. Gene-specific specifications were developed using the MANE Select transcripts NM_000518.5 (*HBB*) and NM_000517.6 (*HBA2*).

Because the molecular consequences of predicted LoF variants depend on transcript context, the VCEP developed gene-specific PVS1 decision trees for *HBB* and *HBA2* to calibrate PVS1 strength based on nonsense-mediated decay (NMD), alternative splicing, and translation re-initiation (Figure 2). Both specifications follow SVI recommendations for NMD prediction, whereby premature termination codons located within the 3′-most exon or the 3′-most 50 nucleotides of the penultimate exon are not expected to trigger NMD [30].

**Figure 2.**
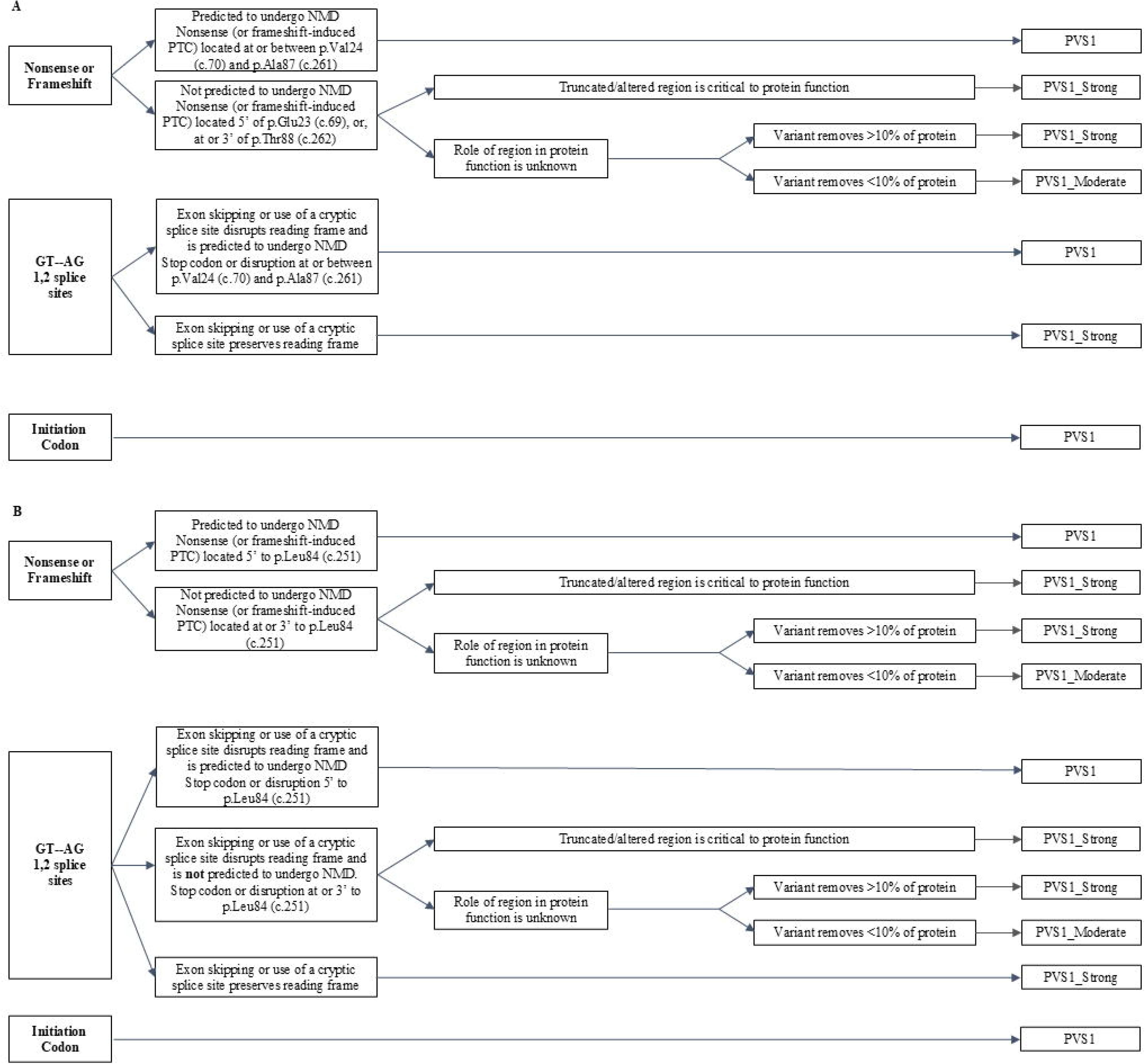
PVS1 specifications. Hemoglobinopathy VCEP PVS1 decision trees for predicted loss-of-function variants in *HBB* (A) and *HBA2* (B), adapted from the ClinGen SVI recommendations[30]. PVS1 assessments were performed using the MANE Select transcripts NM_000518.5 (*HBB*) and NM_000517.6 (*HBA2*), both comprising three coding exons. The HBB-specific framework further incorporates published experimental evidence demonstrating NMD escape and translation re-initiation in the 5′ region of exon 1[32].

For *HBB*, published functional studies demonstrating translation re-initiation and NMD escape informed refinement of the decision tree [31–33]. Premature termination codons occurring at or before p.Glu23 (up to c.69) were considered within the 5′ NMD-resistant region, those occurring between p.Val24 (c.70) and p.Ala87 (c.261) were considered NMD-sensitive, and those occurring at or beyond p.Thr88 (from c.262 onward) were evaluated as NMD-resistant with consideration of the predicted production of truncated β-globin chains. In contrast, for *HBA2*, where the NMD boundaries remain less well characterized because of the predominance of deletional disease mechanisms and the limited number of experimentally studied truncating variants [34,35], the standard SVI framework was adopted. Accordingly, premature termination codons occurring before p.Leu84 (c.251) were considered NMD-sensitive, whereas those occurring at or beyond p.Leu84 (c.251), corresponding to the terminal 50 nucleotides of exon 2 and the entirety of exon 3, were predicted to escape NMD.

For both genes, canonical ±1 ±2 splice site variants and initiation codon variants were interpreted based on their predicted transcript consequences, including their expected effects on NMD. These gene-specific considerations informed the assignment of PVS1_Very Strong, PVS1_Strong, and PVS1_Moderate evidence strengths within the VCEP framework.

#### PM4: protein length changing variants

PM4 was retained without modification and applied to in-frame deletions/insertions and stop-loss variants that disrupt protein function. A notable example is the α-globin antitermination variant NM_000517.6:c.427T>C (TAA>CAA, Hb Constant Spring) in *HBA2* exon 3 that accounts for the majority of non-deletional forms of α-thalassemia worldwide [4].

#### PM1: Located in a mutational hotspot and/or critical functional domain

PM1 was retained at the Moderate strength level and specified for regions with established functional importance and limited benign variation. For *HBB*, this included the core promoter TATAA box (c.-80 to c.-76) and the polyadenylation signal sequence (AATAAA), both of which represent recognized mutational hotspots associated with impaired gene expression. For *HBA2*, PM1 was applied to the polyadenylation signal sequence and residues directly involved in binding α-hemoglobin stabilizing protein (AHSP; α32R, α104H, α118F, and α120P), whose interaction is essential for α-globin stability, preventing precipitation of nascent α-globin chains before hemoglobin assembly [8]. Variants affecting these regions disrupt well-established mechanisms regulating globin production or stability. The resulting globin chain imbalance promotes accumulation and precipitation of unmatched globin chains, leading to oxidative membrane damage and ineffective erythropoiesis that characterizes disease pathogenesis in thalassemia syndromes.

#### PM5: Novel variant at a residue or nucleotide position with a previously established pathogenic variant

PM5 was applied when a novel variant occurred at an amino acid residue where a different missense variant had previously been established as pathogenic. In thalassemia, this criterion was additionally extended to non-coding nucleotide positions where a different pathogenic substitution had previously been reported, reflecting the important contribution of regulatory and splicing variants to globin gene disease mechanisms. A caveat to PM5 application is that nucleotide substitutions may alter RNA splicing in addition to changing the encoded amino acid. Therefore, PM5 is not applied when the pathogenicity of the comparator variant is attributable primarily to altered splicing rather than the affected amino acid residue or nucleotide position itself. Such evidence can be assessed independently under PP3/BP4.

#### PS1: Same amino acid change as a previously established pathogenic variant

PS1 was retained according to the original ACMG/AMP recommendations and applied when a variant resulted in the same amino acid substitution as a previously established pathogenic variant, irrespective of the underlying nucleotide change. For *HBA2*, an additional Moderate-strength application (PS1_Moderate) was specified when the same amino acid substitution had previously been established as pathogenic in the paralogous *HBA1* gene. This modification was supported by the high degree of sequence and functional conservation between the two α-globin genes [36].

### Computational data (PP3, BP4, BP7)

Computational evidence thresholds were informed by a comprehensive benchmarking study using 1,627 curated *HBB, HBA2*, and *HBA1* variants and 31 in silico prediction tools. Predictor performance was evaluated and calibrated within the ACMG/AMP Bayesian framework [28] to derive evidence-based thresholds for pathogenic and benign computational evidence. The resulting recommendations identified REVEL as the preferred predictor for missense variants, SpliceAI for splice prediction, CADD PHRED score for non-missense variants, and GERP++ for conservation assessment, and formed the basis for the VCEP specifications of PP3, BP4, and BP7 described below. Further methodological details are provided elsewhere [18].

#### PP3: computational evidence supporting pathogenicity

PP3 was specified using variant type-specific thresholds. For missense variants, PP3 was applied when REVEL >0.8 or SpliceAI Δ score (DS) >0.3; if REVEL was unavailable, CADD PHRED >23.5 was used instead. For non-coding variants, synonymous variants, in-frame insertions/deletions, and stop-loss variants, PP3 was applied when CADD PHRED >12 or SpliceAI DS >0.3. PP3 was not applied to canonical ±1 and ±2 splice site variants or other variants evaluated under PVS1, to avoid double counting of the same evidence.

#### BP4: computational evidence supporting a benign effect

For missense variants, BP4 required REVEL <0.7 together with SpliceAI DS ≤0.3; if REVEL was unavailable, CADD PHRED ≤20 was substituted. For non-coding variants, synonymous variants, in-frame insertions/deletions, and stop-loss variants, BP4 required CADD PHRED ≤11 and SpliceAI DS ≤0.3. Splice prediction was applied to non-canonical splice-altering variants, including those predicted to create cryptic splice sites or activate nearby alternative splice sites.

*BP7: synonymous variants with no predicted functional impact*.

The VCEP adopted BP7 for synonymous variants predicted to have no impact on splicing (SpliceAI DS ≤0.3) and lacking evolutionary conservation (GERP++ <0). BP7 provides additional evidence that a synonymous variant has a low likelihood of being disease-causing and may be applied together with BP4 when the respective requirements for both criteria are met.

### Case level/segregation data (PS2, PS4, PM3, PM6, PP1, BP2, BP5, BS2, BS4)

Case-level observations constitute a major source of evidence for variant interpretation, contributing to multiple ACMG/AMP criteria including PS2/PM6 (de novo occurrence with or without parental testing), PM3/BP2 (allelic observations in trans or in cis), PP1/BS4 (co-segregation in affected family members, or lack thereof), PS4/BS2 (variant observation in cases or controls), and BP5 (alternate locus observations). A distinctive feature of thalassemia is that heterozygous carriers are typically clinically asymptomatic yet exhibit characteristic hematological abnormalities and hemoglobin profiles detectable by routine laboratory testing [37]. Coupled with the long-standing implementation of carrier and newborn screening programs in many regions [38], this has enabled diagnostic laboratories to accumulate extensive case-level evidence for both common and rare globin variants. Importantly, many rare variants are observed only in heterozygous carriers or compound heterozygous individuals and may otherwise remain difficult to classify, making such observations a particularly valuable source of evidence for variant interpretation. The VCEP therefore developed disease-specific specifications to systematically utilize this unique body of evidence, incorporating published observations, curated case-level genotype-phenotype data from IthaPhen [39], and anonymized unpublished data contributed by participating expert laboratories under the ClinGen framework for protected case-level data sharing while safeguarding sensitive genetic and personal health information.

#### PS4: allelic observations in the heterozygous state

The original ACMG/AMP framework permits application of PS4 when the prevalence of a variant is significantly increased in affected individuals compared with controls. However, conventional case-control studies are unavailable for most rare globin variants. To address this limitation, the VCEP developed a phenotype-based point system that quantifies evidence from unrelated heterozygous individuals exhibiting laboratory findings characteristic of thalassemia (Table 3), consistent with recommendations implemented by other VCEPs [40,41]. The scoring system assigns points according to the specificity of the observed phenotype for the relevant gene-disease pair, with cumulative point totals corresponding to evidence strengths of Supporting (0.5–1.49 points), Moderate (1.5–3.49 points), Strong (3.5–15.99 points), and Very Strong (≥16 points). To preserve phenotypic specificity, individual evidence categories were assigned maximum cumulative point contributions, preventing repeated observations of less specific phenotypes from outweighing fewer observations with greater diagnostic specificity.

**Table 3.**
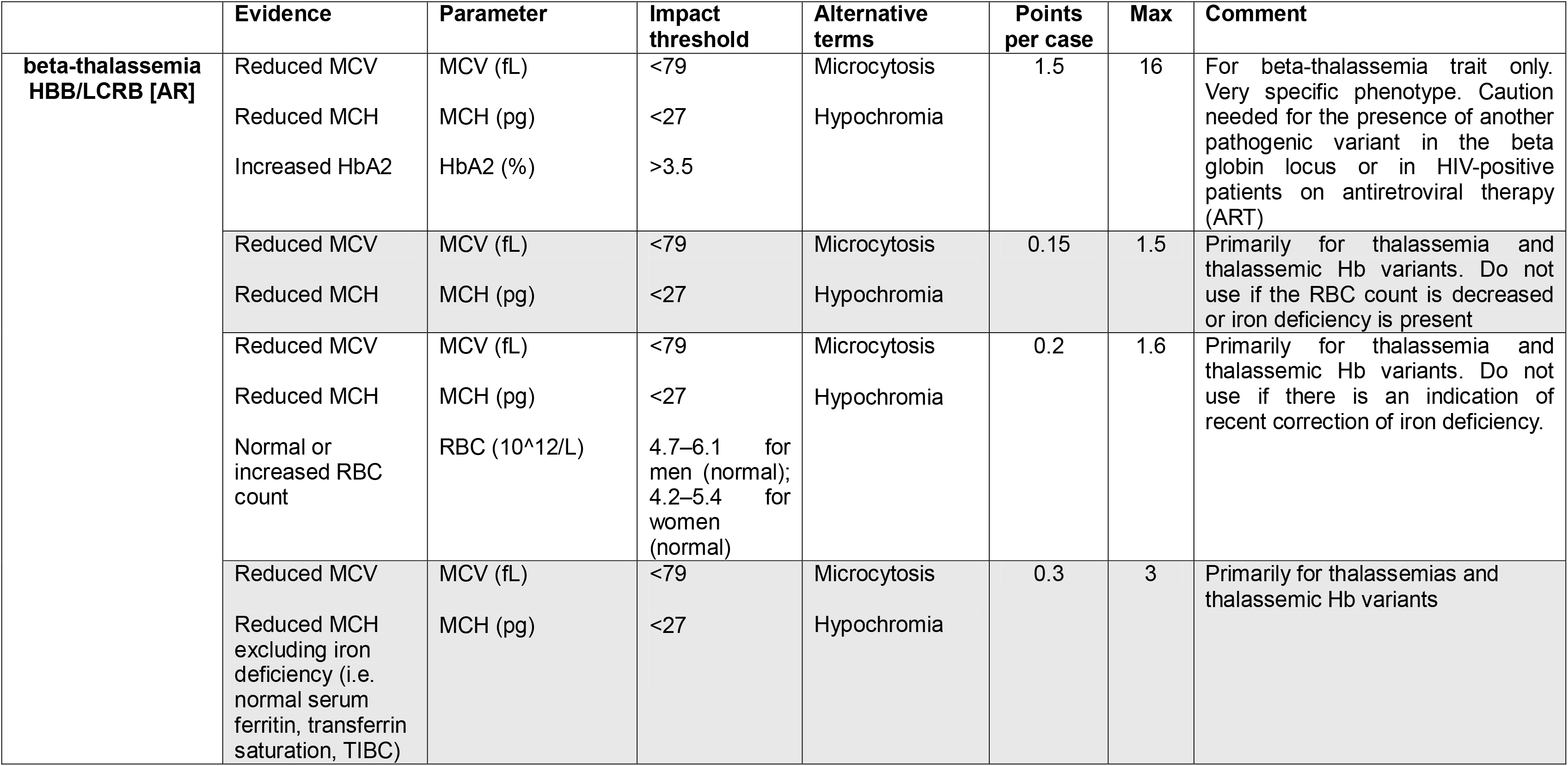
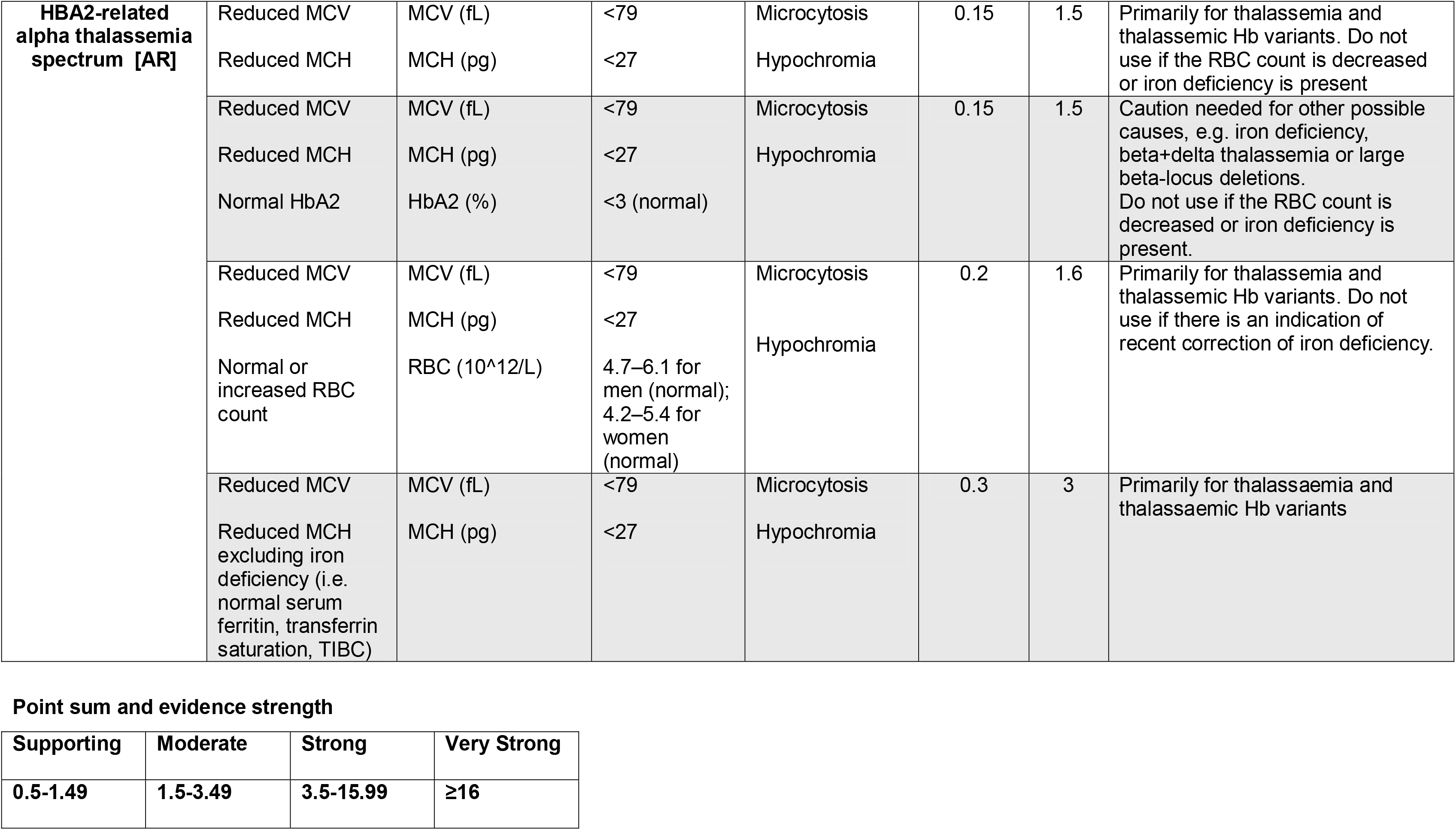
Hemoglobinopathy VCEP point-based framework for evaluating phenotypes in heterozygotes under PS4. Phenotypic and hematological parameters used to score independent heterozygous cases for HBB– β-thalassemia HBB/LCRB and HBA2– α-thalassemia are shown with their respective impact thresholds, points per case, and maximum cumulative scores. Only independent (unrelated) cases are considered; multiple affected individuals within the same family are counted as one case. Where multiple phenotypic evidence levels are met by the same case, only the highest applicable point level is assigned. MCV, mean corpuscular volume; MCH, mean corpuscular hemoglobin; HbA2, hemoglobin A2; RBC, red blood cell count; TIBC, total iron-binding capacity; AR, autosomal recessive.

|  | **Evidence** | **Parameter** | **Impact threshold** | **Alternative terms** | **Points per case** | **Max** | **Comment** |
|---|---|---|---|---|---|---|---|
| **beta-thalassemia HBB/LCRB [AR]** | Reduced MCV<br>Reduced MCH<br>Increased HbA2 | MCV (fL)<br>MCH (pg)<br>HbA2 (%) | <79<br><27<br>>3.5 | Microcytosis<br>Hypochromia | 1.5 | 16 | For beta-thalassemia trait only. Very specific phenotype. Caution needed for the presence of another pathogenic variant in the beta globin locus or in HIV-positive patients on antiretroviral therapy (ART) |
|  | Reduced MCV<br>Reduced MCH | MCV (fL)<br>MCH (pg) | <79<br><27 | Microcytosis<br>Hypochromia | 0.15 | 1.5 | Primarily for thalassemia and thalassemic Hb variants. Do not use if the RBC count is decreased or iron deficiency is present |
|  | Reduced MCV<br>Reduced MCH<br>Normal or increased RBC count | MCV (fL)<br>MCH (pg)<br>RBC (10^12/L) | <79<br><27<br>4.7–6.1 for men (normal); 4.2–5.4 for women (normal) | Microcytosis<br>Hypochromia | 0.2 | 1.6 | Primarily for thalassemia and thalassemic Hb variants. Do not use if there is an indication of recent correction of iron deficiency. |
|  | Reduced MCV<br>Reduced MCH excluding iron deficiency (i.e. normal serum ferritin, transferrin saturation, TIBC) | MCV (fL)<br>MCH (pg) | <79<br><27 | Microcytosis<br>Hypochromia | 0.3 | 3 | Primarily for thalassemias and thalassemic Hb variants |
| **HBA2-related alpha thalassemia spectrum [AR]** | Reduced MCV<br>Reduced MCH | MCV (fL)<br>MCH (pg) | <79<br><27 | Microcytosis<br>Hypochromia | 0.15 | 1.5 | Primarily for thalassemia and thalassemic Hb variants. Do not use if the RBC count is decreased or iron deficiency is present |
|  | Reduced MCV<br>Reduced MCH<br>Normal HbA2 | MCV (fL)<br>MCH (pg)<br>HbA2 (%) | <79<br><27<br><3 (normal) | Microcytosis<br>Hypochromia | 0.15 | 1.5 | Caution needed for other possible causes, e.g. iron deficiency, beta+delta thalassemia or large beta-locus deletions.<br>Do not use if the RBC count is decreased or iron deficiency is present. |
|  | Reduced MCV<br>Reduced MCH<br>Normal or increased RBC count | MCV (fL)<br>MCH (pg)<br>RBC (10^12/L) | <79<br><27<br>4.7–6.1 for men (normal); 4.2–5.4 for women (normal) | Microcytosis<br>Hypochromia | 0.2 | 1.6 | Primarily for thalassemia and thalassemic Hb variants. Do not use if there is an indication of recent correction of iron deficiency. |
|  | Reduced MCV<br>Reduced MCH excluding iron deficiency (i.e. normal serum ferritin, transferrin saturation, TIBC) | MCV (fL)<br>MCH (pg) | <79<br><27 | Microcytosis<br>Hypochromia | 0.3 | 3 | Primarily for thalassaemia and thalassaemic Hb variants |

**Point sum and evidence strength**
**Supporting** **Moderate** **Strong** **Very Strong**
**0.5-1.49** **1.5-3.49** **3.5-15.99** **≥16**

For *HBB–*β-thalassemia, the combination of reduced mean corpuscular volume (MCV <79 fL), reduced mean corpuscular hemoglobin (MCH <27 pg), and elevated HbA₂ (>3.5%) was considered highly specific for the carrier state and therefore assigned the highest per-case point value. In contrast, isolated microcytosis and hypochromia are less specific and receive proportionally lower scores. For *HBA2–*α-thalassemia, hematological findings overlap substantially with iron deficiency and other conditions; therefore, lower point values were assigned unless additional evidence, such as exclusion of iron deficiency or corroborating laboratory parameters, increased diagnostic specificity. Only unrelated individuals were counted, multiple phenotype categories were not combined for the same observation, and potential complex genotype interactions were considered during evidence evaluation.

#### PM3: detected in trans with a pathogenic variant

The VCEP adopted the ClinGen SVI recommendations for PM3 (v1.0), implementing the established quantitative framework in which cumulative observations of a variant in trans with a pathogenic or likely pathogenic allele determine the final evidence strength (i.e., Supporting, Moderate, Strong, Very Strong). Disease-specific specifications were introduced to define qualifying phenotypes and acceptable comparator variants for thalassemia. Specifically, PM3 evidence was considered only for homozygous or compound heterozygous individuals exhibiting a phenotype more severe than the expected heterozygous carrier state based on hematological and/or clinical findings; when detailed laboratory data were unavailable, a well-established clinical diagnosis (e.g., β-thalassemia major or HbH disease) was accepted as evidence of affected status. To avoid circularity, the pathogenicity of the variant on the opposite allele was required to be established independently of the variant under assessment using VCEP classifications (CSpec) or published expert consensus recommendations [18]. In accordance with the SVI recommendations, both alleles were required to satisfy rarity requirements, homozygous observations were capped at a maximum cumulative contribution of 1.0 point, and repeated observations involving the same recurrent founder pathogenic allele were interpreted cautiously, as they may reflect founder allele frequency rather than independent evidence.

#### PP1/BS4: Segregation with disease or lack thereof

The original ACMG/AMP framework recognizes co-segregation with disease as evidence supporting pathogenicity and allows increasing evidence strength with increasing numbers of informative segregations but does not define quantitative thresholds. The Hemoglobinopathy VCEP therefore adopted the ClinGen SVI likelihood-based approach for autosomal recessive disorders, consistent with recommendations implemented by other VCEPs [42–44], applying PP1_Supporting, PP1_Moderate, and PP1_Strong at LOD scores of >0.9, >1.5, and >2.1, respectively (Table 4). These thresholds correspond to increasing statistical support that the variant segregates with disease within a family, whereas BS4 provides strong evidence against pathogenicity when convincing lack of co-segregation is demonstrated. Disease-specific specifications focused on ensuring that only informative segregations, as defined under the ClinGen framework [17], contributed to evidence assessment. Accordingly, the same criteria used to define affected status for PM3 were applied when evaluating segregation evidence. In addition, compound heterozygous individuals contributed to segregation analyses only when the two variants were confirmed to be in trans, and caution was exercised in families harboring additional potentially pathogenic variants or established genetic modifiers that could confound interpretation.

**Table 4.**
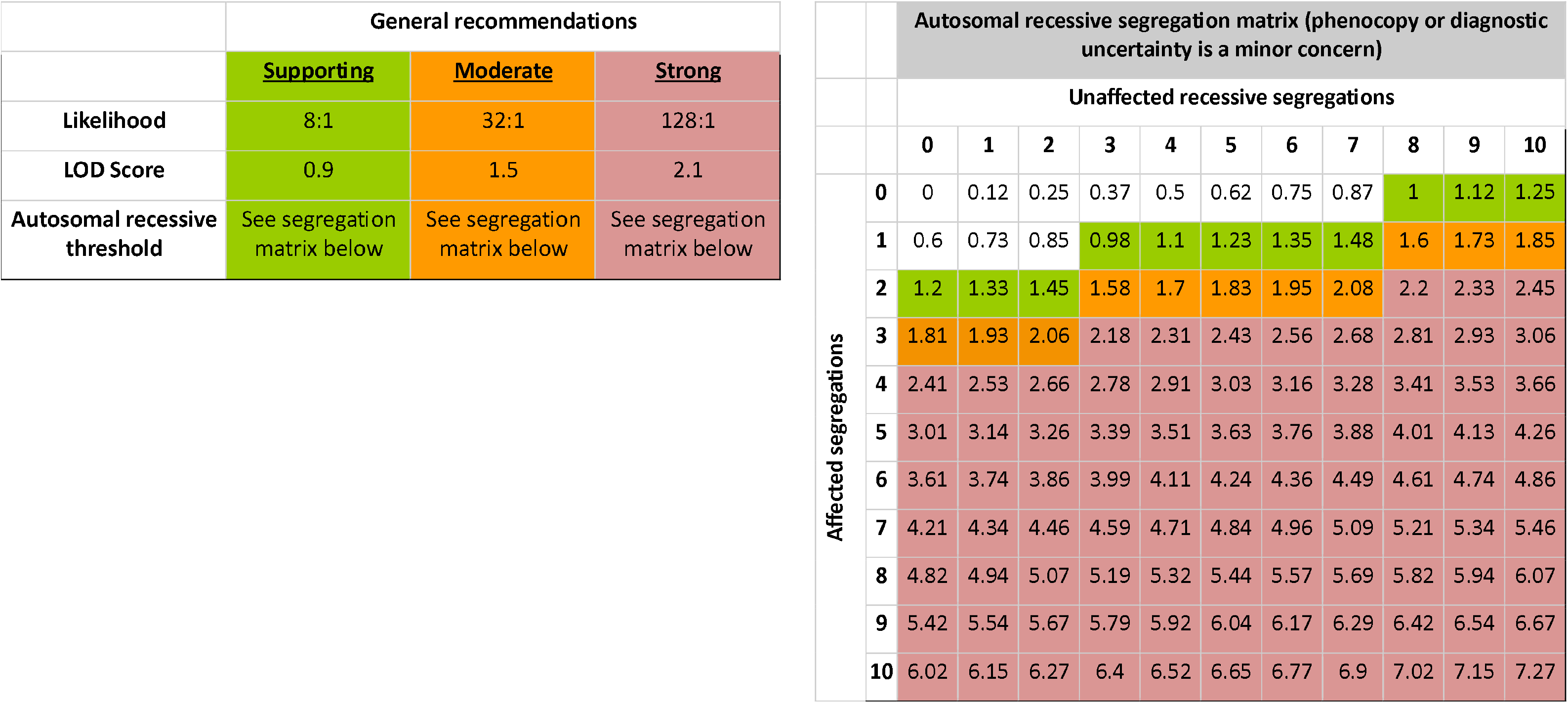
Hemoglobinopathy VCEP recommendations for PP1 segregation evidence. PP1 evidence strength is assigned according to the calculated logarithm of the odds (LOD) score, with thresholds of >0.9, >1.5, and >2.1 for Supporting, Moderate, and Strong evidence, respectively. LOD scores for autosomal recessive segregation analysis based on the number of affected (rows) and unaffected (columns) informative segregations. Affected segregations include family members in whom the homozygous or compound heterozygous disease genotype co-segregates with the phenotype or who are obligate carriers inferred from informative pedigree structure. Compound heterozygous individuals are counted only when the two variants were confirmed in trans. Unaffected segregations include at-risk relatives (typically siblings) who are wild-type or heterozygous carriers for one familial variant and do not exhibit the disease phenotype; unaffected carrier parents are not considered informative. Individuals lacking either genotype or phenotype information are excluded. Although siblings represent the most common informative relatives, other family members may contribute in specific pedigree structures (e.g., families with multiple affected branches). Each cell indicates the corresponding LOD score[17].

|  | General recommendations |  |  |  |  |  |  |  |  |  |  |  |
|  | Supporting | Moderate | Strong |  |  |  |  |  |  |  |  |  |
|---|---|---|---|---|---|---|---|---|---|---|---|---|
| Likelihood | 8:1 | 32:1 | 128:1 |  |  |  |  |  |  |  |  |  |
| LOD Score | 0.9 | 1.5 | 2.1 |  |  |  |  |  |  |  |  |  |
| Autosomal recessive threshold | See segregation matrix below | See segregation matrix below | See segregation matrix below |  |  |  |  |  |  |  |  |  |
|  |  | Autosomal recessive segregation matrix (phenocopy or diagnostic uncertainty is a minor concern) |  |  |  |  |  |  |  |  |  |  |
|  |  | Unaffected recessive segregations |  |  |  |  |  |  |  |  |  |  |
|  |  | 0 | 1 | 2 | 3 | 4 | 5 | 6 | 7 | 8 | 9 | 10 |
| Affected segregations | 0 | 0 | 0.12 | 0.25 | 0.37 | 0.5 | 0.62 | 0.75 | 0.87 | 1 | 1.12 | 1.25 |
|  | 1 | 0.6 | 0.73 | 0.85 | 0.98 | 1.1 | 1.23 | 1.35 | 1.48 | 1.6 | 1.73 | 1.85 |
|  | 2 | 1.2 | 1.33 | 1.45 | 1.58 | 1.7 | 1.83 | 1.95 | 2.08 | 2.2 | 2.33 | 2.45 |
|  | 3 | 1.81 | 1.93 | 2.06 | 2.18 | 2.31 | 2.43 | 2.56 | 2.68 | 2.81 | 2.93 | 3.06 |
|  | 4 | 2.41 | 2.53 | 2.66 | 2.78 | 2.91 | 3.03 | 3.16 | 3.28 | 3.41 | 3.53 | 3.66 |
|  | 5 | 3.01 | 3.14 | 3.26 | 3.39 | 3.51 | 3.63 | 3.76 | 3.88 | 4.01 | 4.13 | 4.26 |
|  | 6 | 3.61 | 3.74 | 3.86 | 3.99 | 4.11 | 4.24 | 4.36 | 4.49 | 4.61 | 4.74 | 4.86 |
|  | 7 | 4.21 | 4.34 | 4.46 | 4.59 | 4.71 | 4.84 | 4.96 | 5.09 | 5.21 | 5.34 | 5.46 |
|  | 8 | 4.82 | 4.94 | 5.07 | 5.19 | 5.32 | 5.44 | 5.57 | 5.69 | 5.82 | 5.94 | 6.07 |
|  | 9 | 5.42 | 5.54 | 5.67 | 5.79 | 5.92 | 6.04 | 6.17 | 6.29 | 6.42 | 6.54 | 6.67 |
|  | 10 | 6.02 | 6.15 | 6.27 | 6.4 | 6.52 | 6.65 | 6.77 | 6.9 | 7.02 | 7.15 | 7.27 |

#### BS2: observed in unaffected individuals

The original ACMG/AMP framework applies BS2 when a variant is observed in healthy individuals despite full penetrance being expected at an early age. Recognizing the recessive nature of thalassemia, the Hemoglobinopathy VCEP adapted this criterion to observations in homozygous or compound heterozygous individuals who remain asymptomatic or exhibit only the expected heterozygous trait phenotype. A single informative observation met BS2_Supporting, whereas two or more independent observations met BS2. Unaffected individuals were required to be well characterized clinically and hematologically, including a full blood count and hemoglobin characterization, to exclude subtle disease manifestations and identify additional variants or abnormal hemoglobin quantities. In compound heterozygous individuals, the variant on the opposite allele was required to be independently established as pathogenic, and alternative genetic explanations, including pathogenic variants in the corresponding globin genes (i.e., *HBB* or *HBA1/HBA2*, as appropriate) and established disease modifiers, were excluded. Finally, only individuals older than 2 years of age were considered informative, as persistence of fetal hemoglobin during early childhood may attenuate the expected phenotype.

#### BP2: observed in cis with a pathogenic variant

The VCEP retained the original ACMG/AMP definition of BP2, applying the criterion when the variant under assessment was observed in cis with an independently established pathogenic variant. The VCEP did not apply BP2 if the phenotype was more severe than expected for either variant in isolation or if the variant under assessment had only ever been reported in cis with a pathogenic variant, as its significance in isolation could not be established. Pathogenicity of the accompanying variant was required to be established using VCEP classifications (CSpec) or published expert consensus recommendations.

#### PS2/PM6: de novo occurrence

The VCEP retained the original ACMG/AMP definitions of PS2 and PM6, distinguishing confirmed de novo occurrences (PS2) from assumed de novo occurrences in which biological parentage was not verified (PM6). For hemoglobinopathies, these criteria were considered applicable in individuals with disease as well as those exhibiting a characteristic heterozygous trait phenotype (Table 3), provided there was no family history consistent with inheritance. Furthermore, PS2 was used only when no other established pathogenic variant was present that could explain the observed phenotype.

#### BP5: alternate molecular explanation

The VCEP applied and specified BP5 only for *HBB*–β-thalassemia. The criterion was met when the observed phenotype could be fully explained by an alternative molecular basis independent of the variant under assessment. Representative examples include β-thalassemia intermedia resulting from co-inheritance of an α-globin locus duplication and a correspondingly aggravated imbalance of the α-globin/β-globin ratio [45], or β-thalassemia-like phenotypes caused by pathogenic variants in *SUPT5H* [46]. In such circumstances, the alternative molecular diagnosis provides a more plausible explanation for the phenotype, supporting a benign interpretation of the variant under evaluation.

### Functional data (PS3, BS3)

#### PS3_Supporting, BS3_Supporting: functional evidence

Functional studies provide direct evidence of the biological consequences of genetic variants and can therefore contribute to pathogenic or benign classification when they assess mechanisms relevant to disease pathogenesis. For thalassemia, the VCEP reviewed available functional assays and specified those that directly interrogate established disease mechanisms, including impaired globin gene expression, abnormal RNA splicing, defective globin chain synthesis, and structural or stability defects of variant hemoglobins. Although many of the approved assays are well established and routinely used in the diagnosis and characterization of hemoglobinopathies, the published literature does not consistently report the standardized validation controls and performance characteristics required to support higher evidence strengths under the ClinGen SVI framework [47]. Moreover, interpretation of some assay readouts, particularly transcript-based assays, may require consideration of the downstream molecular consequences of the observed effect. Accordingly, the VCEP specified these assays at the Supporting evidence level (PS3_Supporting and BS3_Supporting). Assay-specific recommendations are summarized in Supplementary Table 2.

##### Globin biosynthetic assays

Thalassemias are characterized by an imbalance in the normal coordinated synthesis of the α– and β-globin chains (1:1 ratio) that make up the hemoglobin tetramer. Accordingly, assays that quantify globin chain synthesis and evaluate the α:β synthesis ratio, directly assess the primary molecular defect underlying these disorders and provide functional evidence of impaired globin production [48,49].

**Hemoglobin stability assays**: Disruption of the normal structure of the hemoglobin molecule may reduce globin stability, resulting in intracellular precipitation and accelerated degradation of the affected globin chain [8]. The contribution of globin instability to thalassemia pathogenesis varies according to the underlying molecular defect. For example, elongated α-globin variants such as Hb Constant Spring (c.427T>C) and Hb Paksé (c.429A>T) in *HBA2* produce unstable proteins that contribute directly to the thalassemic phenotype, whereas variants such as Hb E (c.79G>A) and Hb Knossos (c.82G>T) in *HBB* primarily act through aberrant splicing and reduced β-globin synthesis but also exhibit altered structural properties, including reduced stability [11,50]. In both settings, reduced globin stability may decrease the effective pool of functional globin available for hemoglobin assembly and exacerbate α:β chain imbalance. The VCEP distinguishes these variants from classical unstable hemoglobinopathies, in which globin instability is the primary pathogenic mechanism leading to hemolytic anemia, and from dominant β-thalassemia caused by NMD-escaping *HBB* variants that produce highly unstable truncated β-globin chains [51]. Accordingly, hemoglobin stability assays (e.g., heat stability or isopropanol precipitation tests) were considered relevant functional studies for demonstrating a variant-induced reduction in globin stability, when such an effect is consistent with the molecular consequence of the specific variant.

##### Hemoglobin electrophoresis and chromatographic quantification

Hemoglobin electrophoresis, capillary electrophoresis, and high-performance liquid chromatography (HPLC) are routinely used to detect and characterize variant hemoglobins. Within the VCEP specifications, these assays were considered informative only when they demonstrated an abnormal quantity of the variant hemoglobin (e.g., reduced peak or abundance), consistent with impaired globin synthesis, stability, or expression. Detection of a variant hemoglobin based solely on altered electrophoretic mobility or chromatographic retention time was not considered sufficient functional evidence. Furthermore, measurements of hemoglobin fractions such as HbA_2_, HbF or Hb H were not used as PS3_Supporting evidence, as these findings are incorporated into phenotype-based criteria (e.g., PS4, PM3, and PP1) and would otherwise constitute double counting of the same evidence.

##### In vitro splicing assay

Aberrant RNA splicing is an established disease mechanism underlying many forms of α– and β-thalassemia, with pathogenic variants disrupting normal pre-mRNA processing through canonical splice-site alterations or activation of cryptic splice sites [52]. Accordingly, in vitro splicing assays were considered relevant functional studies for variants with potential effects on RNA processing. These assays evaluate the splicing pattern generated by the variant, for example using minigene plasmid-based constructs or transcript analyses, and determine whether the variant produces aberrant splice products, including cryptic splice-site activation, partial intronic inclusion, or exon skipping [53,54]. Detection of aberrant splice products, including in the presence of residual wild-type transcript, was considered evidence that the variant disrupts normal RNA processing, whereas exclusive detection of the expected wild-type transcript supported normal splicing. Importantly, although detection of an abnormal transcript provides direct evidence that a variant disrupts RNA splicing, its downstream consequence should not be inferred from the splicing assay alone. Rather, interpretation should consider whether the aberrant transcript is predicted to undergo NMD, preserve the reading frame, or produce an altered protein product, as these features may influence the application of other ACMG/AMP criteria, including PVS1 and PM4 [47].

##### In vitro-cell based assay

Reporter assays and transfection studies in erythroid cell models (e.g., K562, HEL, or HUDEP-2 cells) were considered relevant functional studies when they directly evaluated the effect of a variant on globin gene expression. These assays typically involve transfection with constructs carrying mutant promoter or enhancer sequences linked to a reporter cassette, or expression of cloned mutant globin genes, followed by quantification of reporter activity or globin mRNA/protein levels relative to the corresponding wild-type construct. Because reduced globin expression is an established disease mechanism in thalassemia, experimentally demonstrated decreases in transcription or expression provide direct evidence of variant impact on gene function. The VCEP considered such assays appropriate when they specifically assessed the molecular consequence of the variant under investigation using suitable wild-type controls, biological replicates, and statistics. This specification also encompasses RNA studies performed in cells derived from heterozygous or homozygous individuals, where assessment of wild-type and aberrantly transcripts can provide direct evidence of variant effects on gene expression or RNA processing.

### Pilot application of the VCEP specifications

To evaluate the applicability of the gene-/disease-specific criteria, the final specifications were piloted independently on 20 *HBB* and 13 *HBA2* variants. Variants with established ClinVar classifications, primarily supported by a two-star review status, were selected to assess concordance with classifications derived using the VCEP specifications. The pilot set also included two VUS, one for each gene. The ClinVar and final VCEP classifications, criteria applied, and supporting evidence summaries for each pilot variant are provided in Supplementary Table 3.

Following application of the respective specifications, 16/20 (80%) *HBB* variants (Figure 3A) and 11/13 (84.6%) *HBA2* variants (Figure 3B) were concordant with their existing ClinVar classifications at the exact or broader P/LP or B/LB level. Overall, 27 of 33 variants (81.8%) were concordant. The two VUS included in the pilot remained unresolved. Four additional variants―one *HBA2* variant classified as B, two *HBB* variants classified as LB, and one *HBB* variant classified as P/LP in ClinVar―were classified as VUS due to conflicting evidence identified during curation. Across both pilot sets, the final VCEP classifications comprised 17 P, two LP, six VUS, one LB, and seven B variants, with no reclassification opposing pathogenic and benign classification categories. In an exploratory analysis, classification using the Bayesian framework described by Tavtigian et al [28] was concordant with the VCEP classification for 32/33 (97.0%) variants. The only discordant result involved the *HBB* variant classified as P/LP in ClinVar but resolved as VUS by the VCEP; it was classified as P under the Bayesian framework, illustrating how quantitative integration of conflicting evidence may resolve classifications that remain uncertain under the categorical ACMG/AMP framework.

**Figure 3.**
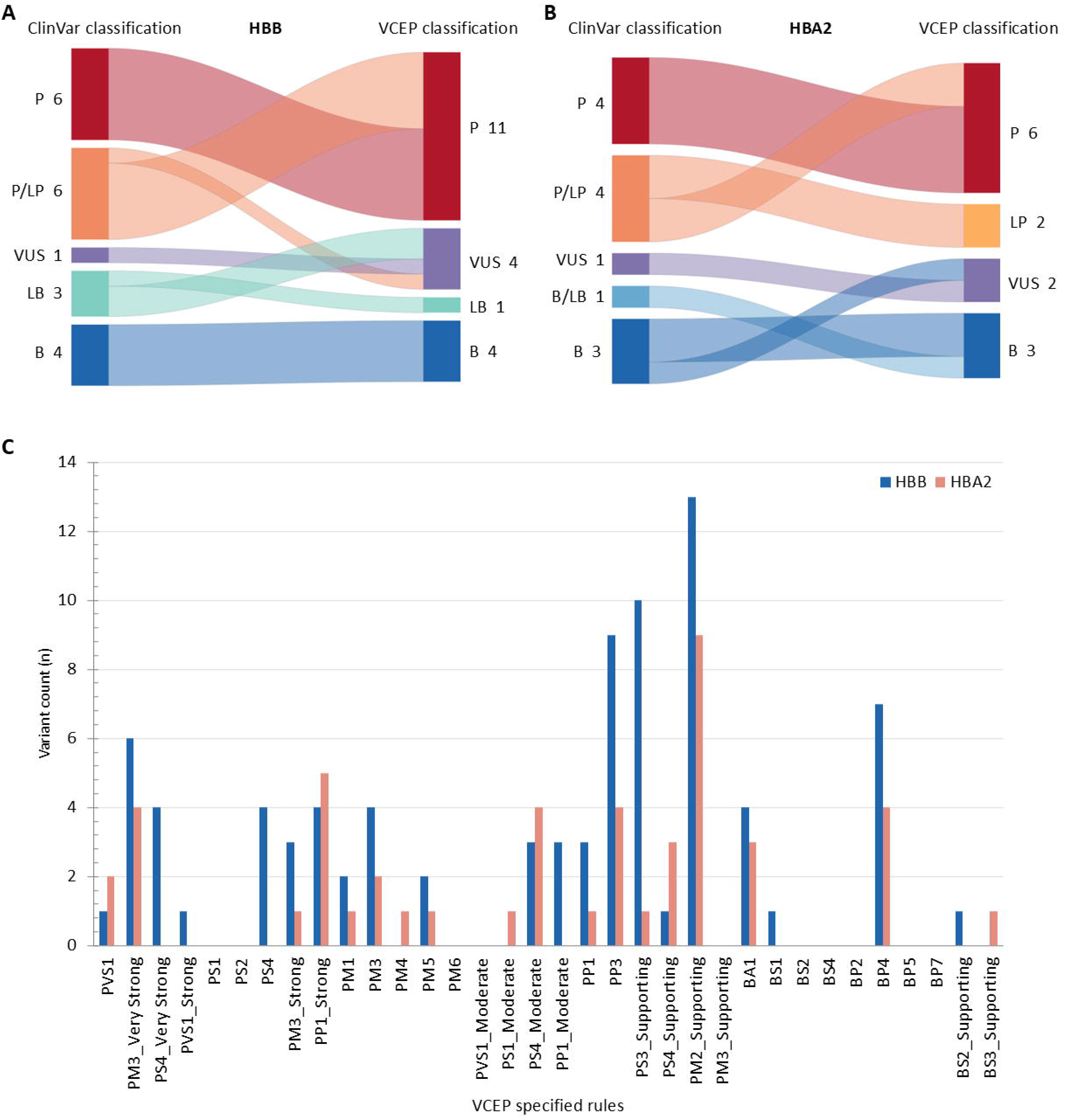
Pilot application of the Hemoglobinopathy VCEP gene– and disease-specific ACMG/AMP criteria. Sankey diagrams illustrate classification outcomes for **(A)** *HBB* and **(B)** *HBA2* pilot variants. ClinVar classifications are shown on the left and final VCEP classifications on the right. Colors denote classification categories; flow widths are proportional to the number of variants. **(C)** Frequency of individual ACMG/AMP criteria applied across the *HBB* and *HBA2* pilot variant sets. Bars indicate the number of variants for which each criterion was applied, stratified by gene. P, Pathogenic; LP, Likely pathogenic; VUS, variant of uncertain significance; LB, Likely benign; B, Benign.

The pilot sets collectively enabled application of a broad range of evidence criteria, spanning population data (BA1, BS1, PM2_Supporting), variant type and location (PVS1 at Strong and Very Strong strengths; PS1_Moderate; PM1; PM4; PM5), computational data (PP3, BP4), case-level/segregation data (PS4 at Supporting to Very Strong strengths; PM3 at Moderate to Very Strong strengths; BS2_Supporting; PP1 at Supporting to Strong strengths), and functional data (PS3_Supporting, BS3_Supporting) (Figure 3C). Overall, 20/24 pathogenic and 5/10 benign criterion-strength specifications were represented in the pilot, with representation across all evidence strength levels, including Stand-Alone evidence. Criteria not encountered largely reflected the molecular basis and recessive inheritance of beta-thalassemia HBB/LCRB and HBA2-related alpha thalassemia spectrum, including the rarity of de novo occurrence (PS2, PM6) and the predominance of variants that reduce or abolish globin chain synthesis rather than impact the globin amino sequence, limiting opportunities to apply criteria relevant to amino acid substitution (PS1) or synonymous variants (BP7) [11]. Other criteria required observations not represented in the selected pilot set, including lack of segregation (BS4; captured under PP1), in cis co-occurrence (BP2), or an alternative molecular basis for disease (BP5).

## Discussion

These ClinGen-approved specifications represent an important milestone for the hemoglobinopathy community by systematically translating decades of accumulated molecular, clinical, and laboratory knowledge into disease-specific ACMG/AMP criteria [55,56]. Rather than redefining existing knowledge, they codify well-established principles of hemoglobinopathy diagnosis into a transparent framework that can be applied consistently across laboratories. The high concordance observed during pilot curation, particularly for community-annotated pathogenic and benign variants, demonstrates that the specifications faithfully capture current understanding of globin variant pathogenicity while providing a rigorous approach for interpreting variants with conflicting or limited evidence. Importantly, uncertainty was largely retained where supporting evidence remained insufficient, emphasizing that the framework standardizes interpretation without implying an unwarranted level of certainty.

The need for such harmonization is becoming increasingly evident as diagnostic testing expands from targeted assays to multigene panels, exome sequencing, and whole-genome sequencing, leading to the identification of an ever-growing number of rare globin variants requiring clinical interpretation [57]. Despite *HBB* being one of the best-characterized Mendelian disease genes, ClinVar (accessed 14 July 2026) continues to contain a considerable number of variants with uncertain (∼300) or conflicting (∼100) classifications. These observations illustrate that extensive biological knowledge alone does not ensure consistent variant interpretation and underscore the need for disease-specific specifications supported by expert review. Since the recognition of ClinGen by the FDA in 2018, ClinGen Variant Curation Expert Panel assertions have been recognized as valid scientific evidence for clinical test development and validation. Collectively, these developments highlight the importance of reliable, transparent, and harmonized variant interpretation for carrier screening, genetic counselling, diagnosis, prognosis, and the clinical management of hemoglobinopathies.

The specification process highlighted several challenges unique to hemoglobinopathies. Perhaps the most notable was the interpretation of population frequency data. Unlike many rare Mendelian disorders, pathogenic globin variants are not necessarily rare in the general population because of the evolutionary history of these disorders, including the phenomenon of heterozygote advantage associated with historical malaria-driven balancing selection [58]. Consequently, population frequency alone cannot be interpreted as evidence against pathogenicity, particularly for variants enriched in specific ancestral populations. Accordingly, BA1, BS1, and PM2_Supporting required disease-specific specification to avoid excluding pathogenic variants on the basis of frequency alone, including specific exemptions for established pathogenic variants with elevated population frequencies [27].

Defining the appropriate disease context for variant interpretation presented another important challenge. Clinical severity in thalassemia is often determined by the combined effects of pathogenic variants within the globin gene loci together with established genetic modifiers, producing a continuous spectrum ranging from clinically silent carrier states to severe transfusion-dependent disease. In addition, the same globin gene may be associated with multiple gene-disease pairs that differ in inheritance pattern, molecular mechanism, and clinical presentation. For example, pathogenic variants in *HBB* may cause recessive β-thalassemia, dominantly inherited β-thalassemia, or structural hemoglobin variants (e.g., Hb E (c.79G>A), Hb C (c.19G>A), and Hb S (c.20A>T)), with the resulting phenotype further influenced by variation at modifier loci, including *HBA1* and *HBA2*. Alternative molecular causes, such as pathogenic variants in *SUPT5H*, may also produce β-thalassemia-like phenotypes. Conversely, pathogenic *HBA2* sequence variants contribute not only to *HBA2*– α-thalassemia but also to the broader α-thalassemia spectrum through interaction with variants in *HBA1* or α-globin deletions. Consequently, variant interpretation required careful consideration of both the relevant gene-disease pair and the underlying disease mechanism rather than the gene alone.

One of the most important conceptual adaptations concerned the role of phenotypic evidence in variant interpretation. Unlike many recessive genetic disorders, heterozygous carriers of thalassemia variants exhibit characteristic hematological abnormalities—including microcytosis, hypochromia, and, in β-thalassemia, elevated HbA_2_—that can provide valuable evidence for variant pathogenicity. The original ACMG/AMP framework applies PS4 to variants enriched in affected individuals relative to controls and considers highly specific phenotypes separately under PP4 [13]. This distinction is less appropriate for recessive forms of thalassemia, where heterozygous carriers are not simply “unaffected controls” and their phenotype may, for some variants, provide the only available case-level evidence for classification. Consequently, the carrier phenotype was incorporated directly into case-level evidence rather than being treated as separate supporting evidence. Accordingly, the Hemoglobinopathy VCEP incorporated rigorous phenotypic requirements into the graded application of PS4 while considering PP4 redundant, thereby avoiding double counting of the same evidence. Importantly, this data is generated routinely through carrier screening programs and diagnostic hematology services worldwide. Unlike many rare genetic disorders, however, much of this evidence remains unpublished because carriers are generally healthy and do not require long-term clinical follow-up. Diagnostic laboratories therefore represent a uniquely valuable source of genotype–phenotype evidence. Broader sharing of anonymized case-level observations through dedicated hemoglobinopathy resources such as IthaPhen [39], together with continued contribution to ClinGen Expert

Panel curation through the VCI [20], will be essential to strengthen the evidence base underpinning future variant interpretation. The specification of PM3 and PS4 at Very Strong strength also necessitated a corresponding modification of the ACMG/AMP evidence-combining rules. Whereas the original framework was developed around a single Very Strong criterion (PVS1), the Hemoglobinopathy VCEP recognized that two independent Very Strong criteria should be considered sufficient to support a pathogenic classification, reflecting the highly specific hematological phenotypes and well-defined disease mechanisms characteristic of the thalassemia syndromes.

Interpretation of functional evidence required careful distinction between assays that directly evaluate variant function and those that describe downstream disease phenotypes. For example, hemoglobin variant detection and quantification inform variant function, whereas red blood cell morphology reflects phenotypic consequence. Consistent with current ClinGen recommendations [15], most published functional studies were applied at Supporting strength because they lacked the validation required to justify stronger evidence levels. Emerging multiplexed assays of variant effect (MAVEs) may help address these limitations; however, current datasets remain limited to selected regions of the globin loci [59], with substantial evidence gaps for most missense, intronic, and other non-coding variants.

The current specifications provide a robust, dynamic framework for variant interpretation, intended to incorporate future insights. Their structured, disease-specific rules and quantitative thresholds may also facilitate future computational implementation, including AI-assisted evidence retrieval, annotation, and application of variant classification criteria, while retaining expert oversight for evidence assessment and final classification [60]. Continued sharing and discussion of high-quality evidence from diagnostic laboratories, together with sustained international Expert Panel curation, will be essential for resolving remaining variants of uncertain significance, refining evidence thresholds, and expanding these recommendations to additional hemoglobinopathy gene-disease pairs. As active recommendations are maintained through the CSpec registry, the Hemoglobinopathy VCEP specifications will continue to evolve as new evidence emerges, ensuring that variant interpretation reflects current scientific and clinical knowledge.

## Conclusions

The ClinGen Hemoglobinopathy VCEP has established the first ClinGen-approved disease-specific ACMG/AMP specifications for the interpretation of sequence variants associated with *HBB*–β-thalassemia and *HBA2*–α-thalassemia. By integrating expert consensus with systematic evaluation of disease-specific molecular, clinical, and laboratory evidence, these recommendations provide a robust and transparent framework for harmonized variant interpretation across diagnostic laboratories. Beyond improving the consistency and reproducibility of variant classification, this work establishes a foundation for extending standardized variant interpretation across the broader spectrum of hemoglobinopathies, ultimately supporting more accurate genetic diagnosis, counselling, and clinical care through international collaboration and sustained expert curation.

## Declarations

### Ethics approval and consent to participate

Not applicable. Only anonymized hematological and clinical case-level data were used to support case-level evidence criteria, in accordance with the ClinGen Variant Curation Expert Panel (VCEP) Protocol and Variant Curation Standard Operating Procedure (SOP).

### Availability of data and materials

The variant curations generated by the Hemoglobinopathy VCEP are publicly available through the ClinGen Evidence Repository (ERepo): https://erepo.genome.network/evrepo/ui/summary/classifications?columns=ep%26values=Hemoglobinopathy%20VCEP%26matchTypes=exact%26matchMode=and%26pgSize=25.

The complete ACMG/AMP criteria specification documents are publicly available through the ClinGen Criteria Specification Registry (CSpec): *HBB*–β-thalassemia HBB/LCRB (MONDO:0013517), https://cspec.genome.network/cspec/ui/svi/doc/GN170; and *HBA2*– HBA2-related α-thalassemia spectrum (MONDO:0100562), https://cspec.genome.network/cspec/ui/svi/doc/GN173.

### Author contributions

**PK** coordinated the study. **CS** drafted the manuscript, performed the data analysis, and prepared the tables and figures. **CS, IF, BHHF, TTK, LN, JS** collected evidence and curated variants using the ClinGen Variant Curation Interface (VCI). **CB, CLH, MK, JTS, JSW, ZP, CWL** reviewed and approved variant classifications. **JER** provided guidance and technical support during the ClinGen specification review and approval process. **All authors** contributed to the development, refinement, and/or implementation of the disease-specific ACMG/AMP rule specifications, critically revised the manuscript, and approved the final version.

## Data Availability

All data generated or analysed during this study are included in the manuscript and its supplementary materials. Variant classifications and supporting evidence are publicly available through ClinVar and the ClinGen Evidence Repository.

## List of abbreviations

**Abbreviation Definition**

ACMG/AMP: American College of Medical Genetics and Genomics/Association for Molecular Pathology
AF: Allele frequency
AHSP: Alpha-hemoglobin stabilizing protein
AI: Artificial intelligence
B: Benign
CADD: Combined Annotation Dependent Depletion
ClinGen: Clinical Genome Resource
CNV: Copy number variant
COST: European Cooperation in Science and Technology
CSpec: Criteria Specification Registry
DS: Delta score
ERepo: ClinGen Evidence Repository
FAF: Filtering allele frequency
FDA: U.S. Food and Drug Administration
GBD: Global Burden of Disease
GERP++: Genomic Evolutionary Rate Profiling
HBA1: Alpha-globin gene 1
HBA2: Alpha-globin gene 2
HBB: Beta-globin gene
HPLC: High-performance liquid chromatography
LB: Likely benign
LoF: Loss of function
LOD: Logarithm of the odds
LP: Likely pathogenic
MANE: Matched Annotation from NCBI and EMBL-EBI
MAVE: Multiplexed assay of variant effect
MCH: Mean corpuscular hemoglobin
MCV: Mean corpuscular volume
MONDO: Mondo Disease Ontology
NMD: Nonsense-mediated mRNA decay
P: Pathogenic
PHRED: Phred-scaled score
RNA: Ribonucleic acid
SNV: Single nucleotide variant
SOP: Standard operating procedure
SVI: Sequence Variant Interpretation
UTR: Untranslated region
VCEP: Variant Curation Expert Panel
VCWG: Variant Classification Working Group
VCI: Variant Curation Interface
VUS: Variant of uncertain significance

## Acknowledgments

This study is based upon work from the HELIOS COST Action (CA22119), supported by the European Cooperation in Science and Technology (COST). This publication was supported in part by the National Human Genome Research Institute of the National Institutes of Health through the following grant: 5U24HG009650-08 and 5U24HG009650-09. The content is solely the responsibility of the authors and does not necessarily represent the official views of the National Institutes of Health. The authors thank Zhiyu Peng and Jun Sun (BGI Genomics, Shenzhen, China) for their contributions to the Step 3 pilot assessment of the Hemoglobinopathy VCEP ACMG/AMP specifications.

## Web resources

ITHANET Portal: https://www.ithanet.eu/

IthaGenes database: https://www.ithanet.eu/db/ithagenes

IthaPhen database: https://www.ithanet.eu/db/ithaphen

IthaMaps database: https://www.ithanet.eu/db/ithamaps

ClinVar: https://www.ncbi.nlm.nih.gov/clinvar/

ClinGen Variant Curation Interface (VCI): https://curation.clinicalgenome.org/

ClinGen Criteria Specification Registry (CSpec): https://cspec.genome.network/

ClinGen Evidence Repository (ERepo): https://erepo.clinicalgenome.org/

ClinGen Gene Curation: https://search.clinicalgenome.org/kb/gene-validity

Genome Aggregation Database (gnomAD): https://gnomad.broadinstitute.org/

## AI Disclosures

ChatGPT (OpenAI) was used to assist with Python-based data processing, data visualization, and language editing. All scientific content, analyses, interpretations, figures, and conclusions were developed, reviewed, and verified by the authors, who take full responsibility for the final manuscript.

