## Supplemental Information for "ClinGen Hemoglobinopathy VCEP ACMG/AMP specifications for *HBB* and *HBA2* variant interpretation in thalassemia"

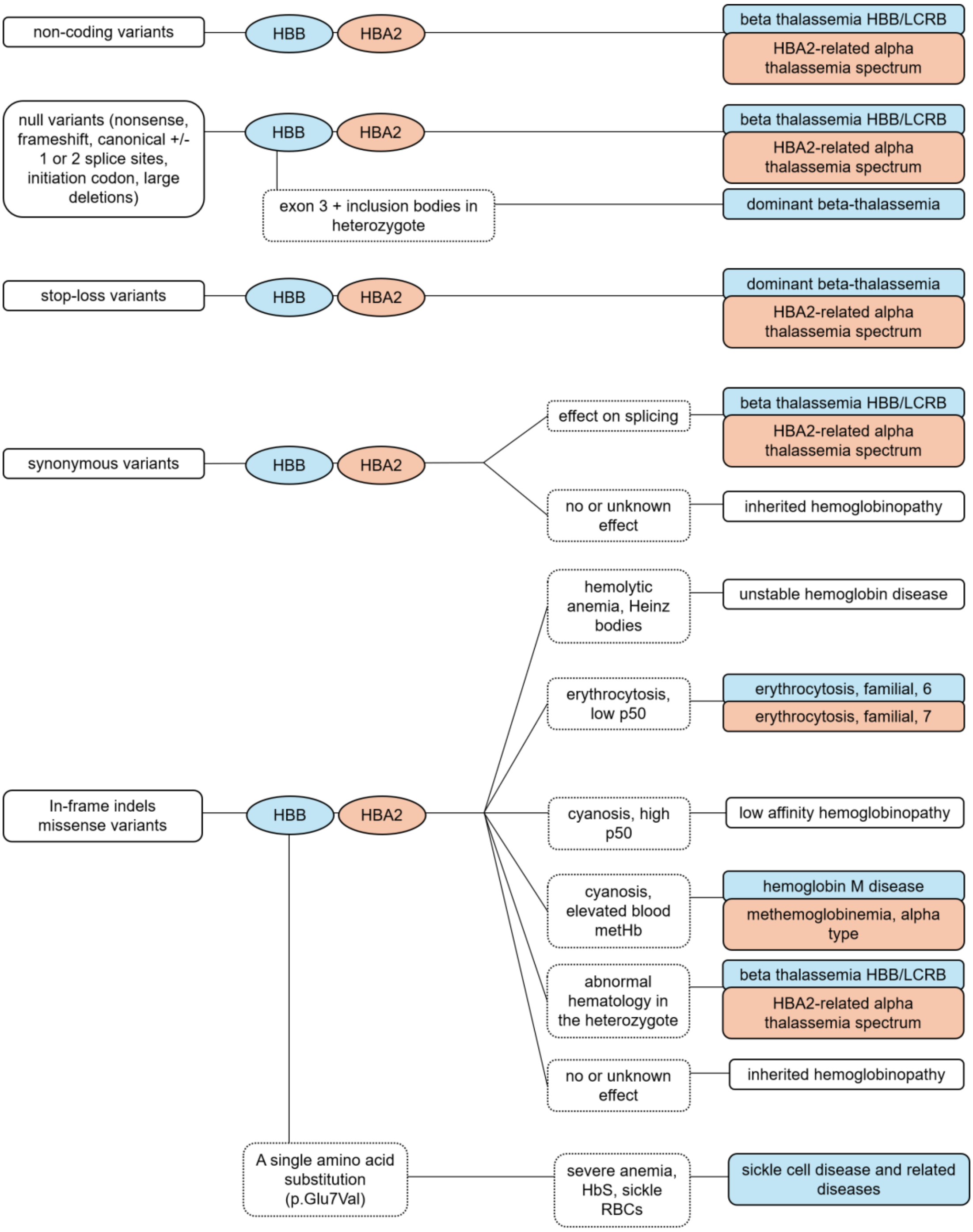


**Supplementary Figure 1. Reference guide for disease term selection during hemoglobinopathy variant curation.** The variant-type-based flowchart was developed to support consistent selection of disease entities associated with HBB and HBA2. Disease terms reflect gene-disease relationships defined through ClinGen gene-disease validity assessment[22] and lumping and splitting recommendations[23]. The Hemoglobinopathy VCEP established the variant type-disease entity associations shown and guides selection of the most appropriate disease entity based primarily on variant type and the availability of evidence supporting pathogenicity. At present, only the *HBB*–β-thalassemia and *HBA2*–α-thalassemia gene–disease pairs are ClinGen-approved for variant classification by the Hemoglobinopathy VCEP; the remaining pathways represent established disease-term associations to support variant curation and future expansion of the VCEP scope. Where available evidence is insufficient to support a specific hemoglobinopathy disease entity, the broader inherited hemoglobinopathy term may be used.

**Supplementary Table 1. *HBB* and *HBA2* exclusion variants.** Variants included in the Hemoglobinopathy VCEP exclusion list are exempt from application of BA1 and BS1 and are not required to meet PM2_Supporting for application of PM3. For each variant, the table provides the HGVS nomenclature, corresponding IthaGenes identifier (ithaID), commonly used variant name, regions of highest reported prevalence, and representative literature supporting their population distribution.

| HGVS name | ithaID | Common name | Regions with high prevalence | PMIDs |
|---|---|---|---|---|
| NM_000518.5(HBB):c.92+5G>C | 107 | IVS I-5 (G>C) | Middle East, South-East Asia | 9576331, 18294253, 12752111, 26865931 |
| NM_000518.5(HBB):c.-79A>G | 25 | -29 (A>G) | Jamaica | 10691857 |
| NM_000518.5(HBB):c.79G>A | 88 | CD 26 GAG>AAG [Glu>Lys] (Hb E) | South-East Asia | 26402558, 24488783 |
| NM_000518.5(HBB):c.92G>C | 100 | CD 30 (G>C) or IVS I (-1) AGG>ACG (Arg>Thr) | India, Pakistan, Maldives | 23162295 |
| NM_000518.5(HBB):c.118C>T | 142 | CD 39 (CAG>TAG) | Mediterranean | 1734721, 21353607, 18096416 |
| NM_000518.5(HBB):c.126_129delCTTT | 147 | CD 41/42 (-TTCT or -CTTT or -TCTT) | South-East Asia | 24534675, 26079343 |
| NM_000518.5(HBB):c.20A>T | 824 | CD 6 GAG>GTG [Glu>Val] (Hb S) | Africa, South-East Asia, Mediterranean | 28361595, 21353607, 26984585 |
| NM_000518.5(HBB):c.19G>A | 810 | CD 6 GAG>AAG [Glu>Lys] (Hb C) | Africa, South-East Asia | 2606477 |
| NM_000518.5(HBB):c.364G>C | 1217 | CD 121 GAA>CA [Glu>Gln] (Hb D-Punjab) | Middle East, South Asia | |
| NM_000518.5(HBB):c.27dupG | 62 | CD 8/9 +G | Middle East | 22460247, 16533735 |
| NM_000518.5(HBB):c.92+1G>A | 101 | IVS I-1 G>A | Mediterranean, Middle East | 21353607, 27199182, 18096416, 29637841 |
| NM_000518.5(HBB):c.93-21G>A | 113 | IVS I-110 G>A | Mediterranean, Middle East | 27199182, 21353607 |
| NM_000518.5(HBB):c.92+6T>C | 111 | IVS I-6 (T>C) | Mediterranean, Middle East | 25408857, 21353607 |
| NM_000517.6(HBA2):c.95+2_95+6delTGAGG | 359 | IVS I-1 (-5 bp) GAGGTGAGG>GAGG----- donor | Mediterranean, Middle East | 26261699, 27199182 |

**ithaID: ID assigned by the IthaGenes database; access using the URL https://www.ithanet.eu/db/ithagenes?ithaID=<ithaID> and substituting the ID number in the table for "<ithaID>".*

**Supplementary Table 2. Functional assays approved by the Hemoglobinopathy VCEP for PS3_Supporting and BS3_Supporting.** Approved assays for *HBB* and *HBA2* variant interpretation, including the biological process assessed, assay readout, evidence thresholds supporting application of PS3 or BS3 at Supporting evidence strength, and important assay-specific limitations. Unless otherwise indicated, the same interpretation framework applies to both genes.

| **Name of assay** | **Biosynthesis assay** | **HPLC / Electrophoresis** | **Hemoglobin stability test** | **In vitro splicing assay** | **Cell-based assay** |
|---|---|---|---|---|---|
| **Measured parameter** | Globin synthesis | Detection and quantification of variant hemoglobins | Hemoglobin stability | Alternative RNA splicing | Gene expression (luciferase/fluorescence, RNA, protein) |
| **Readout description** | Change in biosynthetic ratio of globins | Change in electrophoretic mobilities, or Change in relative peak area and rate of elution (retention time) AND quantification | Reduced stability, precipitation in isopropanol or after heating at 50°C. | Splicing pattern with i) autoradiograms of radiolabeled minigene constructs, ii) long-read RNA sequencing | Transfection of erythroid cell cultures (e.g., K562, HEL, HUDEP-2) with constructs bearing i) reporter cassettes with mutated promoter, 5'UTR and enhancer sequences or ii) a cloned mutant human globin gene. Show changes in gene expression using appropriate methods. |
| **Readout type** | Quantitative | Quantitative | Qualitative | Quantitative | Qualitative / Quantitative |
| **PS3_P evidence (deleterious)** | β-thal trait: β/α 0.50 (0.38-0.62)<br>α0 thal trait: β/α 1.44 (1.22-1.82)<br>HbH: β/α 2.30 (1.80-2.95) | Not concordant with normal chromatogram or readout.<br>HBB: Hb X <35%<br>HBA2: Hb X <15% | Presence of precipitate, visible to the naked eye | Abnormal splice product detected (wild-type and aberrant transcripts are present) | Changes in the expression level of the reporter gene or transgene in comparison to normal levels. |
| **BS3_P evidence (benign)** | β/α 0.96 (0.78-1.14) | Concordant with normal chromatogram or readout | Clear solution (<5% precipitate) | No abnormal splice product detected (only wild-type RNA transcript is present) | No changes in the expression level of the reporter gene or transgene in comparison to normal levels. |
| **Important limitations** | β/α values apply for subjects ≥2 years of age. | Do not apply only for the detection of variant hemoglobins, or for the quantification of normal hemoglobins A, F and A2. | False-positive or inconclusive results may occur when HbF >3% or after prolonged sample storage (methemoglobin formation). Hyper-unstable Hb variants may be rapidly degraded and escape detection; in such cases, no functional evidence should be applied. | Beware of abnormal transcripts that lead to truncated proteins (PS4) without functional consequences, and cell systems where NMD is not active. | Also applies to RNA studies with cells from heterozygous or homozygous probands (wild-type, aberrant transcript detection) |

**Supplementary Table 3. Pilot variant classifications using Hemoglobinopathy VCEP specifications and an exploratory Bayesian framework.** ClinVar classifications and review status (accessed in March 2026) correspond to the most recent ClinVar release available during the final ClinGen VCEP approval process and are presented alongside the final Hemoglobinopathy VCEP classifications and the ACMG/AMP criteria applied. Final VCEP classifications were derived using the ACMG/AMP evidence-combining rules. For comparison, combined odds of pathogenicity, posterior probabilities, and corresponding classifications calculated using the Bayesian framework of Tavtigian et al.(1) are also presented. VUS classifications resulting from insufficient or conflicting evidence are indicated. B, benign; LB, likely benign; VUS, variant of uncertain significance; LP, likely pathogenic; P, pathogenic.

| Variant | ClinVar ID | ClinVar classification | ClinVar review status | VCEP classification | VCEP-specified ACMG/AMP criteria applied | Combined odds of pathogenicity | Posterior probability of pathogenicity | Bayesian classification |
|---|---|---|---|---|---|---|---|---|
| NC_000011.10:g.5225416C>T | 702013 | B | 2 Star | B | BA1, BP4 | 0.48 | 0.050632911 | B |
| NM_000518.5(HBB):c.-75G>C | 36294 | VUS (conflicting evidence) | 1 Star | VUS (insufficient evidence) | PM2_supporting | 2.08 | 0.187725632 | VUS |
| NM_000518.5(HBB):c.92+13G>T | 869245 | LB | 2 Star | VUS (conflicting evidence) | PM2_supporting, BP4 | 0.9984 | 0.099855977 | VUS |
| NM_000518.5(HBB):c.316-185C>T | 36316 | B | 2 Star | B | BA1, BP4 | 0.48 | 0.050632911 | B |
| NM_000518.5(HBB):c.315+81C>T | 439774 | B | 2 Star | B | BA1, BP4 | 0.48 | 0.050632911 | B |
| NM_000518.5(HBB):c.92+5G>C | 15447 | P | 2 Star | P | PP3, PP1_strong, PS3_supporting, PS4, PM3 | 6550.851873 | 0.998628018 | P |
| NM_000518.5(HBB):c.315+260A>G | 750903 | LB | 1 Star | LB | BS2_supporting, BS1 | 0.024 | 0.002659574 | LB |
| NM_000518.5(HBB):c.*111A>G | 15488 | P | 2 Star | P | PM3_very strong, PS4_moderate, PM1, PS3_supporting, PM2_supporting, PP3 | 59051.89542 | 0.999847615 | P |
| NM_000518.5(HBB):c.52A>T (p.Lys18Ter) | 15401 | P | 2 Star | P | PVS1_strong, PM3_very strong, PP1_moderate, PS4, PM2_supporting | 1102306.806 | 0.999991835 | P |
| NM_000518.5(HBB):c.316-36C>T | 799593 | LB | 1 Star | VUS (conflicting evidence) | PM3, PM2_supporting, BP4 | 4.323072 | 0.32448012 | VUS |
| NM_000518.5(HBB):c.316-146T>G | 36312 | P | 2 Star | P | PM2_supporting, PS3_supporting, PP3, PP1, PS4_moderate, PM3_strong | 1515.593879 | 0.994096789 | P |
| NM_000518.5(HBB):c.-137C>G | 15464 | P/LP | 2 Star | P | PM2_supporting, PS4_very strong, PM5, PP1_strong, PM3_very strong, PS3_supporting, PP3 | 89259735.69 | 0.999999899 | P |

| Variant | ClinVar ID | ClinVar classification | ClinVar review status | VCEP classification | VCEP-specified ACMG/AMP criteria applied | Combined odds of pathogenicity | Posterior probability of pathogenicity | Bayesian classification |
|---|---|---|---|---|---|---|---|---|
| NM_000518.5(HBB):c.316-197C>T | 15458 | P | 2 Star | P | PM3_very-strong, PS4_very strong, PP3, PS3_supporting, PM2_supporting, PP1 | 2292922.778 | 0.999996075 | P |
| NM_000518.5(HBB):c.380T>G (p.Val127Gly) | 15483 | P/LP | 2 Star | VUS (conflicting evidence) | PM3_strong, PP1_moderate, PM2_supporting, PS3_supporting, BP4, PS4 | 3144.408899 | 0.997145946 | P |
| NM_000518.5(HBB):c.316-106C>G | 15457 | P/LP | 2 Star | P | PM2_supporting, PS4_very-strong, PM3_very strong, PP3, PS3_supporting, PP1_strong | 20614257.66 | 0.999999563 | P |
| NM_000518.5(HBB):c.82G>T (p.Ala28Ser) | 15239 | P/LP | 2 Star | P | PP1_strong, PS4_moderate, PS3_supporting, PM2_supporting, PP3, PM3 | 3155.058412 | 0.997155552 | P |
| NM_000518.5(HBB):c.114G>A (p.Trp38Ter) | 15405 | P/LP | 2 Star | P | PVS1, PM2_supporting, PP1, PM3, PS4_supporting | 13637.85114 | 0.999340507 | P |
| NM_000518.5(HBB):c.93-21G>A | 15454 | P/LP | 2 Star | P | PP3, PM3_very strong, PP1_moderate, PS4, PS3_supporting | 122609.527 | 0.999926602 | P |
| NM_000518.5(HBB):c.-78A>C | 15470 | P | 2 Star | P | PM2_supporting, PS3_supporting, PM5, PM1, PS4_very strong, PM3_strong, PP3 | 1104270.444 | 0.99999185 | P |
| NM_000518.5(HBB):c.316-373C>A | 695343 | B | 1 Star | B | BA1, BP4 | 0.48 | 0.050632911 | B |
| NM_000517.6(HBA2):c.377T>G (p.Leu126Arg) | 15690 | P/LP | 2 Star | LP | PM2_supporting, PS4_moderate, PP3, PM3 | 81.11524096 | 0.900127882 | LP |
| NC_000016.10:g.173009_173013del | 375746 | P | 2 Star | P | PVS1, PP1_strong, PS4_moderate | 28339.85 | 0.999682527 | P |
| NM_000517.6(HBA2):c.89T>C (p.Leu30Pro) | 15651 | P | 2 Star | P | PM2_supporting, PP1_strong, PS4_moderate, PP3, PM3_very strong | 122609.527 | 0.999926602 | P |
| NC_000016.10:g.172832C>A | 869219 | B | 0 Star | VUS | PM2_supporting, BS3_supporting | 0.9984 | 0.099855977 | VUS |
| NM_000517.6(HBA2):c.-24C>G | 439113 | B | 2 Star | B | BA1, BP4 | 0.48 | 0.050632911 | B |
| NM_000517.6(HBA2):c.*107A>G | 439108 | B | 2 Star | B | BA1, BP4 | 0.48 | 0.050632911 | B |

| Variant | ClinVar ID | ClinVar classification | ClinVar review status | VCEP classification | VCEP-specified ACMG/AMP criteria applied | Combined odds of pathogenicity | Posterior probability of pathogenicity | Bayesian classification |
|---|---|---|---|---|---|---|---|---|
| NM_000517.6(HBA2):c.*98T>C | 439111 | VUS (conflicting evidence) | 1 Star | VUS | BP4, PM2_supporting | 0.9984 | 0.099855977 | VUS |
| NM_000517.6(HBA2):c.*92A>G | 15647 | P/LP | 2 Star | P | PM2_supporting, PM1, PS4_moderate, PP1_strong, PM3_very strong | 255240.025 | 0.99996474 | P |
| NM_000517.6(HBA2):c.179G>A (p.Gly60Asp) | 439112 | P | 2 Star | P | PM2_supporting, PP3, PS4_supporting, PP1_strong, PM3_very strong, PS1_moderate | 255027.8162 | 0.999964711 | P |
| NM_000517.6(HBA2):c.313T>C (p.Cys105Arg) | 804215 | P/LP | 2 Star | LP | PM2_supporting, PM5, PS4_supporting, PM3 | 81.11524096 | 0.900127882 | LP |
| NM_000517.6(HBA2):c.427T>C (p.Ter143Gln) | 15624 | P/LP | 2 Star | P | PM2_supporting, PP3, PM4, PM3_very strong, PP1_strong, PS3_supporting | 255027.8162 | 0.999964711 | P |
| NM_000517.6(HBA2):c.2del (p.Met1fs) | 15692 | P | 2 Star | P | PVS1, PS4_supporting, PM2_supporting, PM3_strong, PP1 | 58897.87904 | 0.999847216 | P |
| NM_000517.6(HBA2):c.95+27C>T | 439773 | B/LB | 2 Star | B | BA1, BP4 | 0.48 | 0.050632911 | B |
